# Aging of the Spine: Characterizing genetic and physiological determinants of spinal curvature

**DOI:** 10.1101/2024.02.27.24303450

**Authors:** Frances M. Wang, J. Graham Ruby, Anurag Sethi, Matthew Veras, Natalie Telis, Eugene Melamud

## Abstract

Increased spinal curvature is one of the most recognizable aging traits in the human population. However, despite high prevalence, the etiology of this condition remains poorly understood. To gain better insight into the physiological, biochemical, and genetic risk factors involved, we developed a novel machine learning method to automatically derive thoracic kyphosis and lumbar lordosis angles from dual-energy X-ray absorptiometry (DXA) scans in the UK Biobank Imaging cohort. In 41,212 participants, we find that on average males and females gain 2.42° kyphotic and 1.48° lordotic angle per decade of life. Increased spinal curvature was strongly associated with decreased muscle mass and bone mineral density. Adiposity had opposing associations, with decreased kyphosis and increased lordosis. To gain further insight into the molecular mechanisms involved, we carried out a genome-wide association study and identified several risk loci associated with both traits. Using Mendelian randomization, we further show that genes fundamental to the maintenance of musculoskeletal function (COL11A1, PTHLH, ETFA, TWIST1) and cellular homeostasis such as RNA transcription and DNA repair (RAD9A, MMS22L, HIF1A, RAB28) are likely involved in increased spinal curvature.

## Introduction

Increased spinal curvature is a frequently observed phenomenon during aging in humans and in other species.^1–3^ Drift to higher thoracic kyphosis and lumbar lordotic angles starts in early ages, is gradual, and takes many decades to develop.^4,5^ These physiological declines are generally considered to be non-pathological unless kyphosis becomes so severe as to exceed more than 40° (hyperkyphosis), or more than 30° in lordosis regions (hyperlordosis).^4^ Hyperkyphosis is known to have a significant heritable component and affects females more than males.^6,7,8^ On the other hand, lumbar hyperlordosis is an understudied condition.^9^

At the extremes, acute injuries, disk fractures, and degenerative joint disorders are believed to be the main drivers of spinal hypercurvature.^10^ In the general population, increases in spine curvature develop largely in the absence of severe events. Prevalent aging conditions such as loss of bone mineral density, decline in muscle strength, and metabolic disorders have been suggested to play an important role.^8,11^ Over time, continued biomechanical stresses on the spine and aging-related declines in the repair capacity of cartilage and bone tissues can lead to the development of vertebral fractures, increased back pain, and consequently reduced mobility.^12,5^ Cumulative adverse consequences are an increased chance of falls, reduced health span, and increased mortality.^13,14^

While the negative consequences of an aging spine on health outcomes are clear, the genetic risk factors and underlying molecular mechanisms of this phenomenon remain poorly understood. Recent advances in the UK Biobank (UKBB) large-scale prospective imaging study create an opportunity to examine changes to spine curvature with age in a general population.^15^ We capitalized on this resource by developing machine learning models to assess spine curvature in an automated fashion across 41,212 dual-energy X-ray absorptiometry (DXA) scans. Using a fully automated machine learning (ML) pipeline, we were able to measure Cobb angles in both thoracic and lumbar regions, thus providing us with an opportunity to examine shared and unique risk factors in different regions of the spine.

Our analysis extends to the study of underlying pathological conditions, physiological, blood biochemistry, and genetic risk factors. We assessed contributions to spine angle from 415 diseases, 15 musculoskeletal traits, 59 biomarkers, and all common genetic variants. We further carry out Mendelian randomization (MR) to test for potential causal involvement of genes within these loci with changes in spinal curvature. To our knowledge, this is the first study to identify genetic risk factors involved in the development of kypholordotic angles.

## Results

### Cobb angle annotation by machine learning

We developed ML models to mimic a well-established Cobb angle measurement technique used in radiological practice.^16^ Briefly: a U-net segmentation model was trained to label the spine body, and a previously published^17^ object-detection model was used to identify appropriate regions for Cobb angle estimation. Training and test data from those models were separate from the Cobb angle preliminary test and validation sets described below.

Cobb angles were estimated between tangent lines drawn through appropriate spine regions using Deming regression. In lateral DXA scan images, human annotators measured two sets of Cobb angles across 320 images: 4 independent annotators measuring T12-T5 kyphotic angle and 3 independent annotators measuring L4-L1 lordotic angle (**Figure 1A**). These images and their annotations were randomly split into a preliminary testing dataset (120 images), used to calibrate parameters of the ML & regression pipeline described above, and a validation dataset (200 images), used for final evaluation of each model’s performance. There was a high degree of concordance across the human annotators (r=0.66-0.91, **Supplementary** Figure 1).

**Figure 1:**
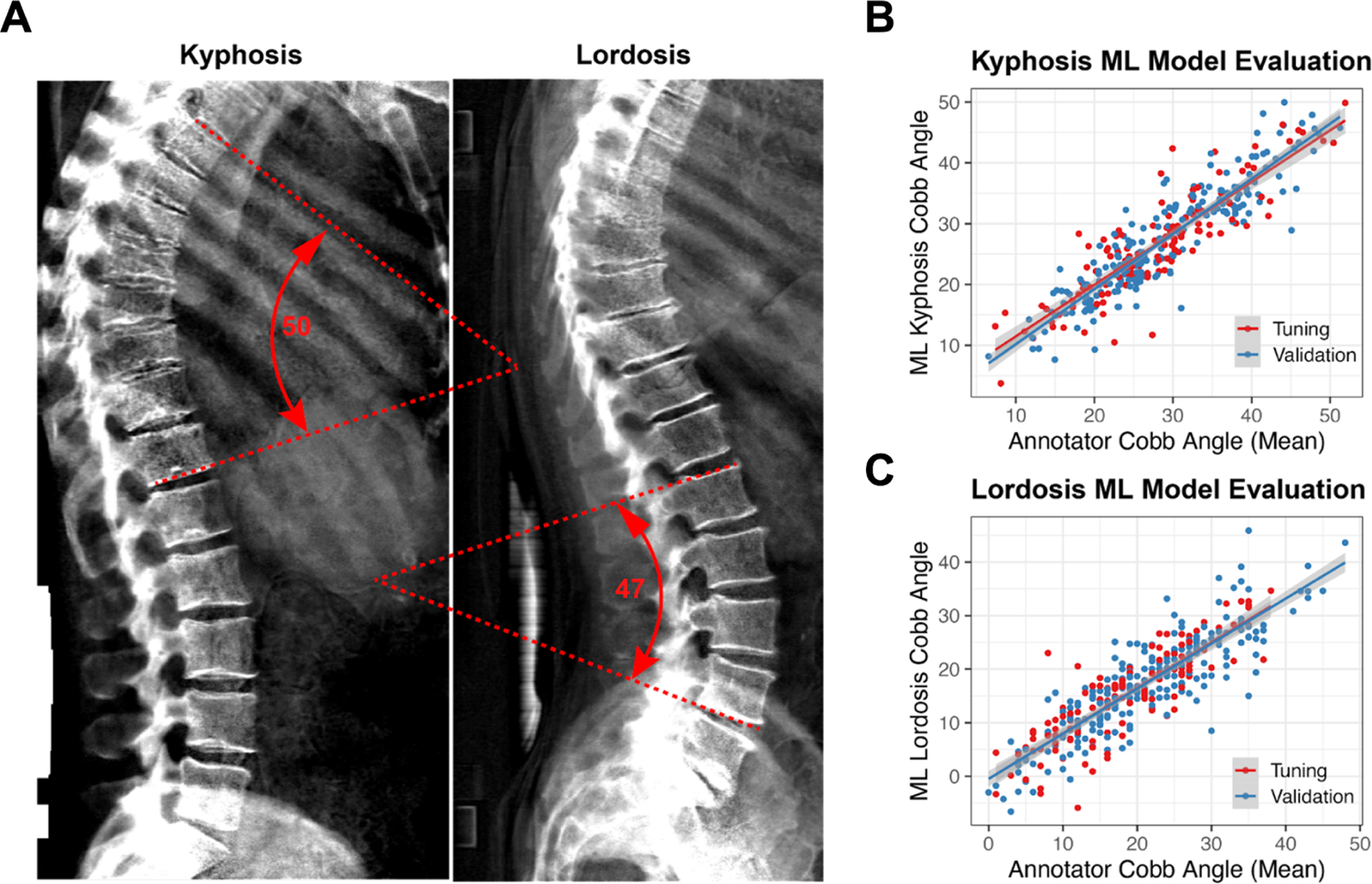
Machine learning model evaluation. A) Example of manually curated kyphotic (T5-T12) and lordotic (L1-L4) Cobb angles in lateral UK Biobank lateral DXA scan. (B-C) Correlations of Cobb angles as assessed by average annotator and machine learning derived kyphotic angles and lordotic angles in training (tuning) and validation sets.

The ML-based estimation algorithms for both kyphosis and lordosis performed well on the 200-image validation dataset, closely matching the results of human annotators with high correlation values (r=0.87 and 0.86 for the kyphosis and lordosis models, respectively) (**Figure 1B and 1C**). The lordosis model has slightly lower accuracy, likely due to greater variability in the ability to define the location of the L4 and L5 junctions.

### Epidemiology of Kyphosis and Lordosis

Among the 41,212 UKBB participants included in this study, 48.3% were male (**Supplementary Table 1**). The median age of DXA imaging subjects was 64 [interquartile interval (IQI) 58, 70] years (**Supplementary Table 1**). A total of 3,702 (9.0%) participants had hyperkyphosis (angle <40°), and 3,109 (7.5%) had hyperlordosis (angle <30°), of which 1,057 (2.6%) participants had both. Thoracic kyphosis Cobb angles were strongly correlated to lumbar lordosis Cobb angles (r=0.49, p-value<0.001, **Supplementary** Fig. 2). Kypholordotic angles were systematically higher in females compared to males at all ages (**Fig. 2A**). Both kyphotic and lordotic Cobb angles increased with age, at approximately the same rate: 2.28° (2.16°, 2.39°) of kyphotic angle and 1.32° (1.19°, 1.44°) of lordotic angle per decade of life (not adjusted for other factors).

**Figure 2.**
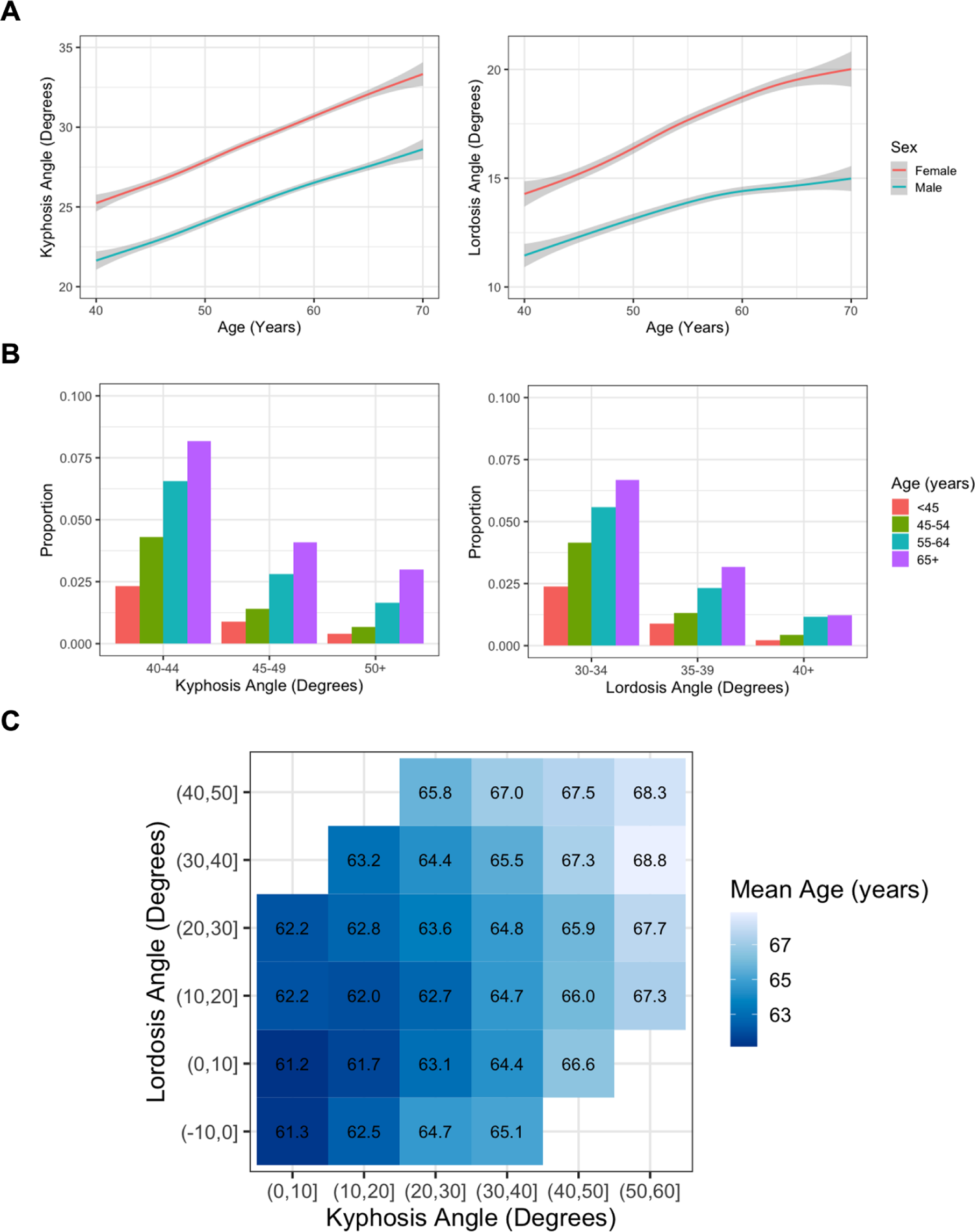
Distribution of kyphosis and lordosis. A) Kyphotic and lordotic angles as a function of age, stratified by sex. Spinal curvature increases with age in both sexes with approximately the same slope, but kyphotic and lordotic Cobb angles are greater in females. B) Prevalence of hyperkyphosis and hyperlordosis increases with age. C) Distribution of kyphotic and lordotic angle by age category and sex.

Since both angles increase with age, we asked if they also increased as a function of each other. In the multivariable model, adjusted for age and sex, the kyphotic angle increased an additional ∼0.4° for every degree of lordotic angle (Supplementary Figure 4). Consequently, with the increase of kypholordotic curvature with age, there was a graded increase in the prevalence of hyperkyphosis and hyperlordosis with age (**Fig. 2B**). By age 75+, the prevalence of hyperkyphosis and hyperlordosis was 15.4% and 10.4%, respectively. Participants with hyperkyphosis were 5.22 (4.89, 5.58) times more likely to also have hyperlordosis than those without hyperkyphosis and, relatedly, participants with hyperlordosis were 4.90 (4.61, 5.21) times more likely to also have hyperkyphosis. Those with higher observed kyphosis and lordosis angles tended to be older (**Fig. 2C**).

### Disease Associations with Spine Curvature

Various pathological pre-existing conditions were associated with increased spine angles (**Fig. 3A**). First, among the strongest associations for kypholordotic angles were previous diagnoses of intervertebral disc disorders such as disc compression disorders (M41, M47, M51, G55) and osteoporosis (M81). For example, prior diagnosis of multiple sclerosis (MS) was highly associated with increased kyphotic angle by 2.24° (0.83°, 3.64°), potentially due to the effect of MS on posture.^18^ We also observed that a variety of gastric conditions including esophagitis, esophageal disease, and gastro-esophageal reflux disease (GERD) [K21, K22, K20] and, perhaps relatedly, diaphragmatic hernia (K44) were associated with increased kyphotic angles, also possibly due to the association of kyphosis with posture, reduced back muscle strength, and increased intra-abdominal pressure.^19^ Interestingly, we observed that metabolic disorders, such as diabetes, gout, and hyperlipidemia (E11, E14, M10, E78), were associated with decreased spine curvature. This effect was generally more pronounced in females, where diabetes was linked to reduction of kyphotic angle by 1.50° (0.71°, 2.30°) versus 0.63° (0.06°, 1.19°) in males (**Supplementary** Fig. 3). The association with these disorders with kypholordotic angles tended to be linear (**Supplementary** Fig. 4). More comorbid conditions were associated with kyphotic angles than lordotic angles, likely in part due to a higher precision in our machine learning-based estimates of kyphotic angles.

**Figure 3.**
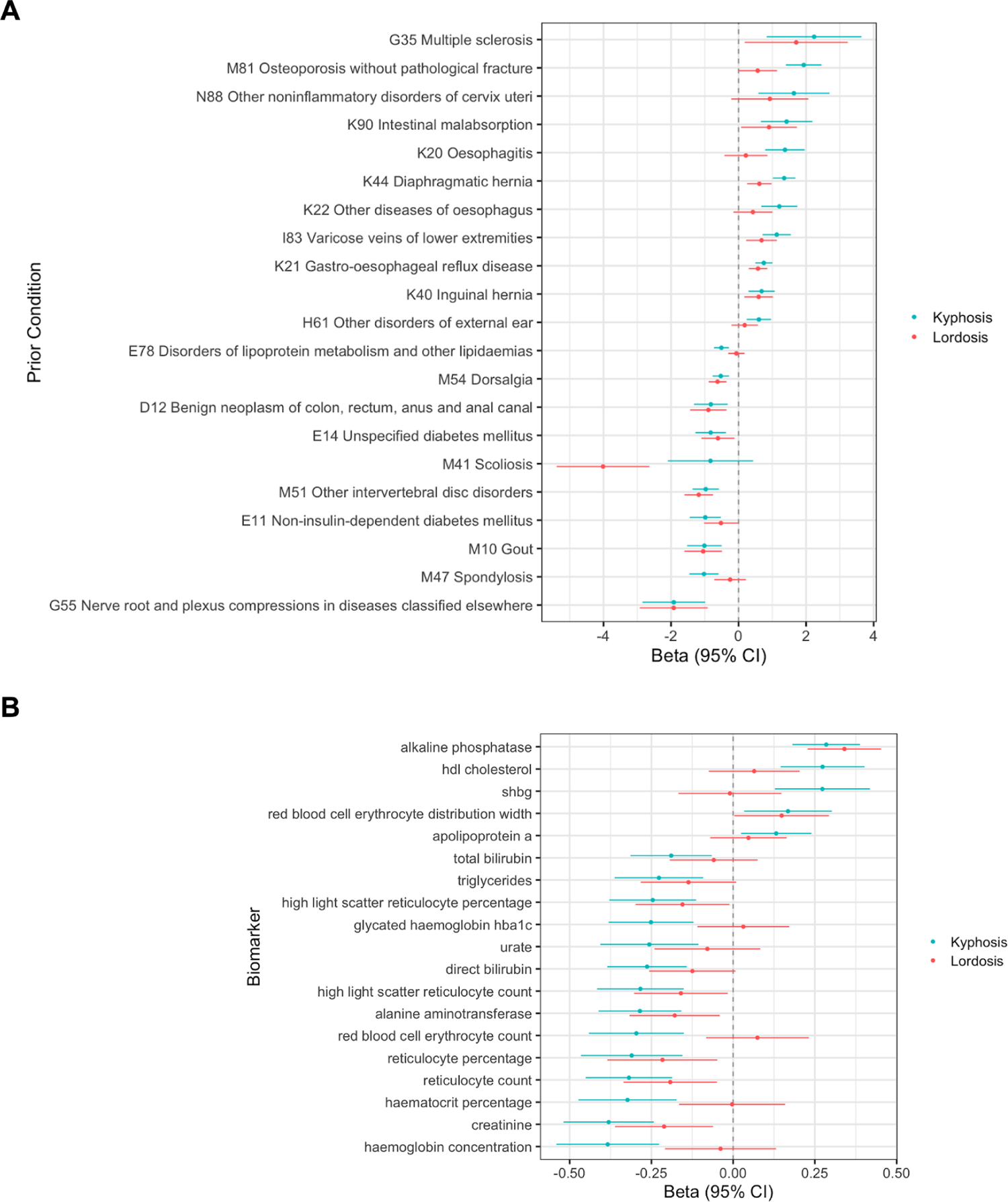
Spinal curvature associations with A) Pre-existing diagnosed conditions B) Biochemistry. All analyses were adjusted by age at visit, sex, BMI, smoking status, and Townsend deprivation index. Angle as a function of a disease or biomarker. Beta is a degree change in kypholordotic angle with the presence of a prior condition or per standard deviation increase in biochemical measure.

In serum biochemistry data, we observed that elevated levels of plasma alkaline phosphatase (ALP) were associated with significant increases in both kyphosis and lordosis angles, supporting established literature that osteoporosis is an important contributing factor to increased spine curvature (**Fig. 3B**). Similar to the inverse associations we found between kypholordotic angles and metabolic comorbidities, in biochemistry data, elevation of metabolic biomarkers (hemoglobin A1C [HbA1c], urate, and triglycerides) were associated with significant reductions in kyphotic angles. This inverse association between biomarkers of metabolic disorders and kyphosis was consistent for both sexes (**Supplementary** Fig. 5).

### Association of Musculoskeletal Traits with Spine Curvature

To evaluate how aging of the musculoskeletal system contributes to increased spinal curvature, we performed a phenotypic association scan with muscle, adiposity, and bone traits across the entire skeletal system in UKBB. Significant associations, adjusted for multiple hypothesis correction, are shown in **Fig. 4A**. We observed that bone mineral density (BMD) measures across the entire skeletal system were strongly associated with spine curvature. Both kyphotic and lordotic angles were consistently larger in those with lower BMD. For example, the kyphotic angle increased by 0.39° (0.31°, 0.47°), and the lordotic angle increased by 0.66° (0.58°, 0.73°) per standard deviation loss of arm BMD. This directional association was mostly consistent for both males and females across skeletal sites, with associations stronger in females than males (**Supplementary** Fig. 6).

**Figure 4:**
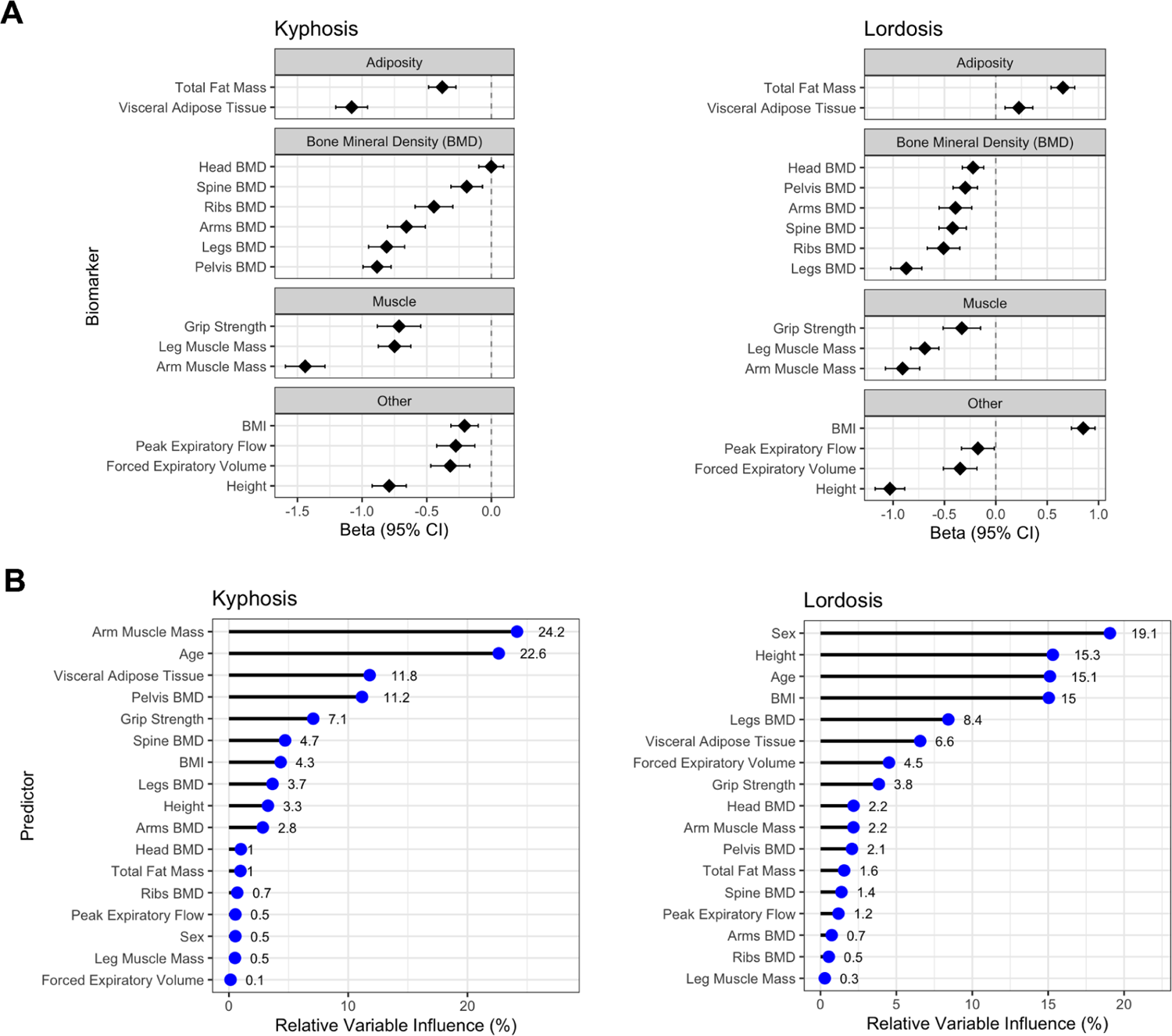
Physiology associations. A) Association of musculoskeletal traits with spine curvature. Analyses were adjusted by age at visit, sex, BMI, smoking status, and Townsend deprivation index, except BMI, height, total fat mass, and visceral adipose tissue were not adjusted for BMI. Beta is a degree change in angle per standard deviation increase in physiological measure. B) Importance of cross-validated GBM-selected variables for predicting spine curvature.

Among muscle measurements, lower muscle mass in both the legs and the arms was also associated with increased spinal curvature, indicating the important role skeletomuscular factors play in the development of pathology (**Fig. 4A**). On average angle increases by 0.91° (0.82°, 0.99°) kyphosis and 1.44° (1.36°, 1.52°) lordosis per standard deviation loss of arm muscle mass (**Supplementary** Fig. 6). Decreased grip strength was also strongly associated with increased spine angles, but association may be due to collinearity with muscle mass.

Interestingly, we observed that fat composition measures showed a differential effect between lordosis and kyphosis. Higher adiposity (total and visceral fat) was linked with increased lordotic angles but reduced kyphotic angles. The same differential effect was observed with body mass index (BMI) and was consistent for males and females (**Supplementary** Fig. 4).

Since physiological measurements are strongly associated and non-linearly dependent on each other, to identify most important predictors of spine curvature we used gradient boosting algorithm (GBM)^20^. While predictors of angles at both sites were similar, their relative contribution (as measured by variable importance) was different (**Fig. 4B**). Consistent with single trait analysis, the top independent predictors of kyphosis were age, muscle mass, BMD, grip strength, and sex. Independent predictors of lordosis were similar to those for kyphosis, but sex, height, and BMI were stronger predictors of lordosis.

### Genetic Risk Factors of Spine Curvature

Leveraging genotyping information, we next evaluated genetic risk factors associated with the development of spinal curvature. We carried out a genome wide association study (GWAS) for both kyphosis and lordosis angles within the participants of Caucasian origin (n=33,413). Both traits had significant heritability h^2^ (kyphosis: 0.34 [0.30, 0.38], lordosis: 0.26 [0.22, 0.30]). We found that there is a strong genetic correlation between kyphotic and lordotic angles (Rg = 0.71, p-value=1.58e-48) indicating that common genetic variation contributes to both traits. The GWASes identified 8 genome-wide significant loci associated with kyphosis and 6 genome-wide significant loci associated with lordosis (**Supplementary** Fig. 9).

Since we observed a strong genetic correlation between kyphosis and lordosis, to increase the statistical power, we carried out multi-trait analysis of GWAS (MTAG) to jointly model these traits.^21^ The results of MTAG were generally consistent with individual trait GWAS (**Fig. 5A**). Given greater power and precision of MTAG, we focus our downstream analyses of the MTAG summary statistics in the remaining sections.

**Figure 5.**
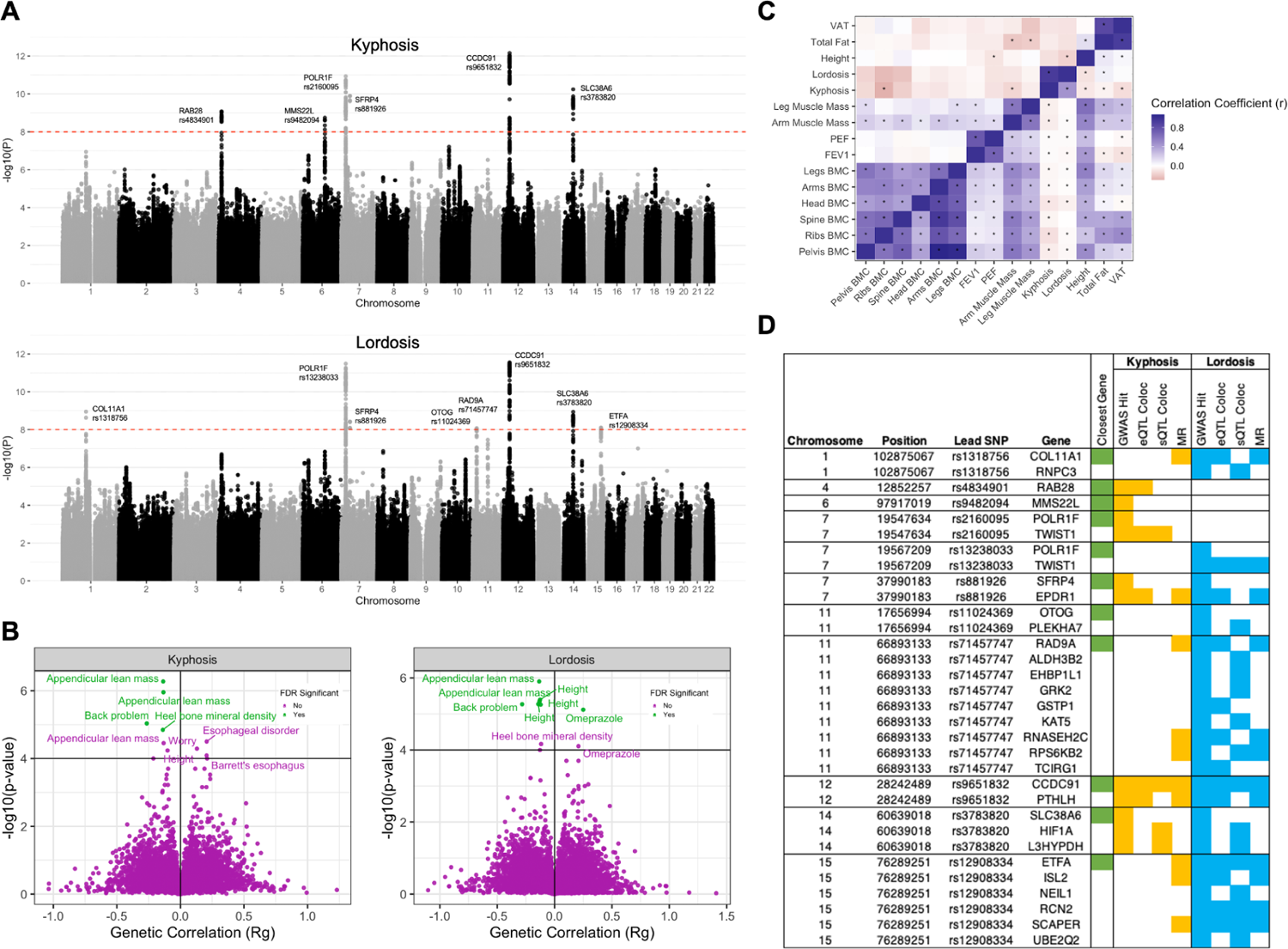
Genetics of spine curvature. A) GWAS manhattan plot with fine-mapped lead SNPs annotated B) GWAS catalog phenotypic correlations with kypholordotic angle. In green are significant associations where pFDR<0.05. C) Comparison of genetic and phenotypic correlations. Displayed in the upper left triangle are genetic correlations for selected musculoskeletal traits. Displayed in the lower right triangle are phenotypic correlations of selected musculoskeletal traits. Both traits show similar patterns of genetic and phenotypic correlations, with greater kypholordotic angles anti-correlated with other traits. Significant correlations are marked with asterisks. D) Genetic summary figure of GWAS, colocalization (eQTL and sQTL), and MR associations by lead SNP and gene.

**Figure 6:**
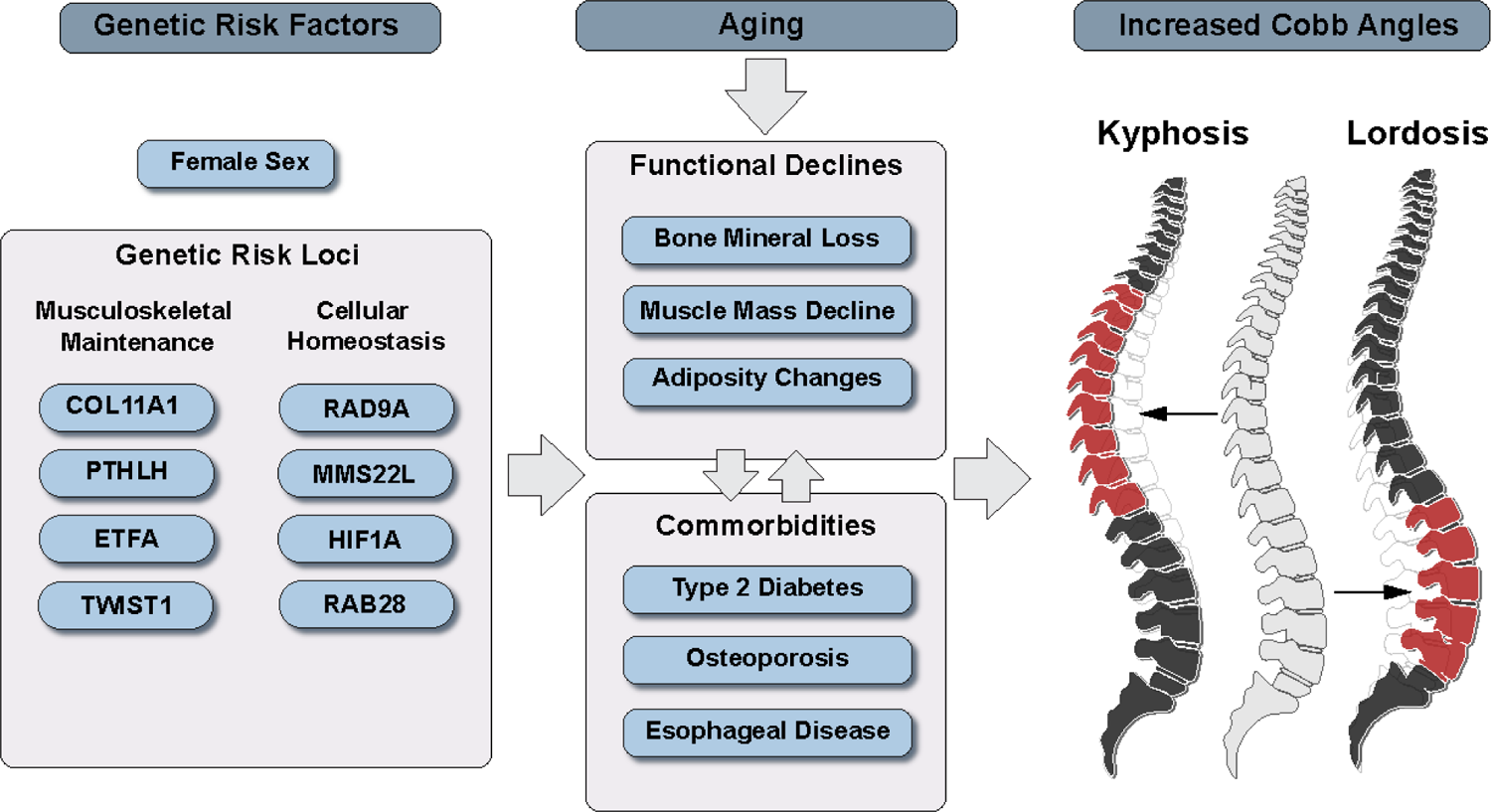
Multi-system physiology effect on spine curvature as a function of age. Loss of bone mineral density, decreased muscle mass, and decreased lung capacity are associated with increased kypholordotic angles. Adiposity has opposing effects on the thoracic and lumbar spine: decreasing kyphosis and increasing lordosis. Aging is a common risk factor for declines in physiological function and presentation of comorbid conditions such as osteoporosis, type 2 diabetes, and esophageal diseases. Both traits are strongly genetically correlated and share common risk alleles such as PTHLH → BMD, COL11A1 → connective tissues, and ETFA → muscle mass, as well as a number of genes involved in DNA repair and cellular homeostasis processes (RAD9A, MMS22L, HIF1A, RAB28).

### Genetic Correlation

To examine if the genetic variation linked to spine angles shares genetics with other traits, we calculated genetic correlations with 10,170 and 10,138 GWAS catalog phenotypes for kyphosis and lordosis, respectively (**Fig. 5B, Supplementary Table 2**). Top traits associated with kyphosis were measures of muscle mass (appendicular lean mass), bone mineral density (heel, total body), and diseases related to the esophagus (esophageal disorder, Barrett’s esophagus, hiatal hernia, GERD). The top genetic correlations of lordosis were similar to that of kyphosis but with stronger genetic associations with body type measures including height, BMI, waist circumference (**Supplementary Table 3**). In line with epidemiological association with GERD diagnosis, we observed a strong genetic correlation of both kyphosis and lordosis with esophageal diseases and omeprazole usage (R_g_ between 0.10-0.25, all p-values<1E-3), further suggesting that esophageal diseases share genetic risk factors with spine curvature traits.

To assess the similarity of demographic-adjusted genetic and phenotypic correlations, we computed an all-by-all correlation matrix for selected musculoskeletal traits and displayed it in a single heatmap (**Fig. 5C**). While most traits showed strong phenotypic correlation with one another, genetic correlations were less statistically powered and therefore more sparse. As expected, strong interdependence between all musculoskeletal traits can be observed with shared genetic origins. A mechanistic understanding of how these traits contribute to the development of spine curvature requires a deeper understanding of the loci involved.

### Gene Prioritization

To better understand genetic risk factors we carried out two types of analysis (i) fine-mapping followed by colocalization with Genotype-Tissue Expression (GTEx) RNA splicing and RNA expression quantitative trait loci (eQTLs) (ii) MR with GTEx RNA expression as mediator of Cobb angle outcomes. Combining these multiple lines of support, we were able to prioritize genes with high likelihood of being involved in development of kypholordotic angles.

Fine-mapping identified 12 unique variants associated with kypholordotic angles (7 with kyphosis and 8 with lordosis) (**Table 1, Supplementary Table 4**). Of these, there were 3 overlapping fine-mapped lead single nucleotide polymorphisms (SNPs) shared by kyphosis and lordosis: rs881926 near SFRP4, rs9651832 near CCDC91, and rs3783820 near SLC38A6. Variation near RAB28 and MMS22L genes showed a stronger association with kyphosis, while variation near COL11A1, OTOG, RAD9A, and ETFA genes showed a stronger association with lordosis. Full fine-mapping results can be found in **Supplementary Table 5 and 6.** Previous GWAS studies have implicated SNPs in these loci with multiple musculoskeletal traits such as height, heel bone mineral density, appendicular lean body mass, various measures of lung function, and spinal stenosis (**Supplementary** Fig. 10).

**Table 1.**
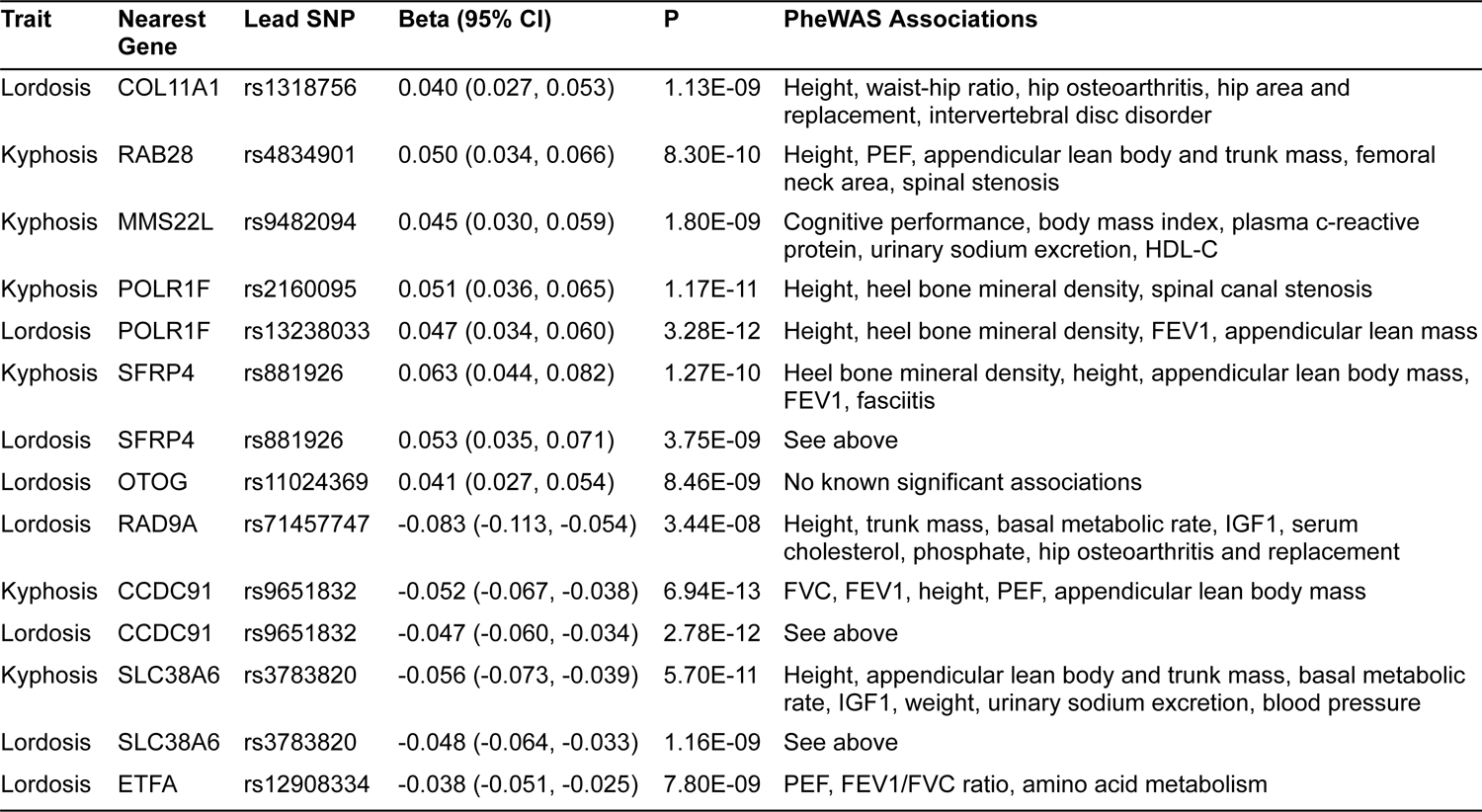
Genetic associations between phenotypic traits and kypholordotic angle.

Using variants identified in fine-mapping with either lordosis or kyphosis, we ran colocalization analysis of Cobb angles with RNA expression data (eQTLs and splicing quantitative trait loci [sQTL]) across multiple tissues profiled in GTEx v8 (**Supplementary Table 7)**. (See Methods). Although, for some genes we observed colocalization with both traits, overall colocalization found more association with lordosis (**Figure 5D**). Some of the fine-mapped SNPs, colocalized with RNA expression of multiple genes within the same loci. For example, we observed colocalization of kyphotic and lordotic angles with both PTHLH and CCDC9 on the chromosome 12 locus, along with both H1F1A and L3HYPDH on chromosome 14.

We followed up colocalization analysis with MR. In MR, genetic variants (instruments) are used to estimate the causal effect of an exposure (RNA expression) on an outcome (spinal curvature). The detailed description of methodology and its limitations can be found in the Methods. Given correlated RNAseq expression patterns observed in colocalization analysis, we can not rule out the possibility of horizontal pleiotropy, but we did find support for the potential mediation effect of multiple genes.

We found that decreased expression of RAD9A (multiple tissues), ETFA (skeletal muscle), COL11A (adipose tissue), ISL2 (esophagus), and increased expression of RNASEH2C (brain), and CCDC91/PTHLH and SCRAPER (multiple tissues) were significantly associated with increased Cobb angles (**Supplementary Table 8 and 9, Fig. 5D**).

Following MR-STROBE guidelines^22^, we evaluated sensitivity of our MR results. We repeated analyses using MR-Egger and median-weighted MR methods, and found that effect sizes were concordant across different estimation methods. MR-Egger didn’t find that associations were statistically significant, but this was expected, given lower statistical power of this methodology. When multiple instruments were available, to avoid estimation bias we tested for heterogeneity of instruments. We did not find significant heterogeneity across instruments.

We summarized our findings in **Figure 5D**. Overall, a complex picture of genetic risk factors emerges, involving genes expressed in multiple tissues with fundamental roles in maintenance of tissues and cellular homeostasis. This included genes such as: COL11A1 (maintenance of connective tissue), RAB28 (intracellular trafficking), MMS22L (DNA double-strand break repair), TWIST1 (transcription factor involved in skeletal-vascular development), EPDR1 (glycoprotein related to neurogenesis), PLEKHA7 (key adherens junction component), RAD9A (DNA double-strand break repair), PTHLH (parathyroid like hormone), HIF1A (hypoxia response including angiogenesis and glycolysis), and ETFA (mitochondrial fatty acid beta-oxidation). Many genes also displayed non-specific tissue expression patterns.

## Discussion

Increased spinal curvature is a universally recognized aging trait; however, the genetic and physiological etiology of this trait has not been well-characterized. In this study, we leveraged machine learning models to automatically quantify kypholordotic angles for 40,224 community-dwelling middle-aged to older adults in UKBB. Physiology, serum biochemistry, and genetics data were analyzed to build an integrated epidemiological and biological understanding of spinal curvature.

We observed that increased curvature in both the lumbar and thoracic spine were strongly intercorrelated and age-dependent (2.4° and 1.5° per decade of life). Previous investigations of the effect of aging on spine angles in males and females did not paint a consistent picture of sex-specific differences in kypholordotic angle and how they change with age.^23,24^ In our analysis, we see a consistent aging effect in both males and females, but with a different baseline of higher kypholordotic angles in females by the age of 40. Although we do not have measurements at earlier ages, likely, significant differences in spine curvature arise much earlier in life or due to developmental differences between sexes. While our cross-sectional analysis of aging effects could be biased by characteristics of the cohort; they are consistent with previous longitudinal assessment of aging effects.^25^

As the population ages, both hyperkyphosis and hyperlordosis increased, reaching a substantial prevalence of 15.4% and 10.4%, respectively, by age 75 in the study. While the prevalence and assessment of hyperkyphosis has been well-characterized, the prevalence of hyperlordosis and its cause is limited.^26,27^ Previous epidemiological studies of hyperkyphosis have implicated several potential pre-existing conditions, such as vertebral fractures and osteoporosis, that could contribute to the development of this pathology.^26^ We have conducted a systematic evaluation of all common pre-existing conditions associated with Cobb angles. The majority of pre-existing conditions associations (multiple sclerosis, osteoporosis, and gastroesophageal diseases) were the same between kyphosis and lordosis, matching in both direction and effect sizes. In our analysis of biochemical factors, we likewise observed similar correspondence between kyphosis and lordosis. These results lead us to conclude that the increase in spinal curvature at two sites is driven by a similar set of underlying factors.

Our analysis of physiological measures suggested that the loss of muscle mass (and relatedly muscle strength) and decreased bone mineral density were the factors most strongly associated with increased spinal curvature. The slope of age-dependent angle increase was approximately the same for both males and females, even though females on average have lower muscle mass, lower BMD, and higher prevalence of osteoporosis. This suggests that the aging effect is not due to baseline differences in BMD and muscle measures and further highlights the important, complex relationship between spinal curvature with muscle mass and BMD with age.^8,28–30^

Examining the effect of body adiposity measures on spine angles, we were surprised to find that increased adiposity (total fat mass, visceral fat, and BMI) was associated with decreased kyphotic angles, but increased lordotic angles. Among physiological measures considered, adiposity was the only measure that showed this opposing relationship between the two traits. This effect remained after adjusting for age, sex, and respective spine angles. The increased lumbar lordosis may be due to the biomechanics of the spine, where the lumbar region compensates for increased adiposity in the trunk by moving the spine forward to adjust the center of gravity.^31^ Alternatively, decreased kyphotic angles were linked to a higher prevalence of metabolic disorders (type 2 diabetes, gout, elevated glucose, and HbA1c), but less so with lordotic angles. It is possible that metabolic disorders can result in an incidental increase in BMD, perhaps strengthening the upper spine and thus decreasing kyphotic angle, while more abdominal adiposity can increase the load on the lower spine.^31–33^

Genetic analyses elucidated the complex origin of spine curvature traits. To our knowledge, this is the first GWAS of kyphosis and lordosis published to-date. We found that angles at both sites have substantial heritability (25-35%) and are strongly genetically correlated, indicating that common genetic drivers influence angles at both sites. Genetic correlation with other traits was consistent with phenotypic associations, with appendicular lean muscle mass and BMD among the strongest correlated traits.

Interestingly, in both phenotypic and genetic associations, we observed an anti-correlation of height with spine angles (higher kyphosis/lordosis associated with smaller stature/height), but it was not clear to us if this was a trivial observation. However, this seems to indicate that there is at least in part shared genetics that determine these skeletally-related traits.

We also observed a strong genetic correlation of increased spine curvature with the development of gastroesophageal disease (GERD and Barrett’s esophagus). This genetic correlation was consistent with increased use of proton-pump inhibitors (PPIs) such as omeprazole and GERD being among the strongest associated pre-existing conditions, as well as future GERD diagnosis events. It is plausible that genetically driven decline in muscle mass is a common factor contributing to both conditions.^34^ Increased incidence of GERD is also known to be associated with metabolic syndrome (eg. diabetes, obesity)^35,36^, but we did not observe a significant genetic correlation between GERD and metabolic syndrome, indicating that metabolic syndrome is unlikely a common genetic risk factor between GERD and spine curvature. The link between these gastroesophageal diseases and spine curvature may also lie in the common use of PPIs for treatment. Chronic use of PPIs which has been associated with increased fracture risk and osteoporosis which may therefore lead to increased spinal curvature.^37,38^

From genetic colocalization and MR, most of the alleles and implicated genes we identified were previously reported in GWASAtlas as associated with broad muscular-skeletal traits such as height, BMD, appendicular lean muscle mass, and maintenance of connective tissues.^39^ Molecular functions and expression patterns of implicated genes did not point to any single cell type or specific tissue involvement, rather they indicated involvement of global cellular maintenance processes. For example, we found support for the hypothesis that expression of genes involved in DNA repair processes (RAD9A), and microtubule-dependent chromosome segregation (SCAPER).^40,41^ Potentially in line with the observation that fundamental DNA repair processes are playing a role, previous studies have demonstrated that overactive p53 activity, and partial loss of mitochondrial checkpoint protein BubR1 can lead to increased senescence and dramatic kypholordosis.^42,43^

A potential causal association of parathyroid hormone (PTH)-like hormone (PTHLH) with the development of kyphosis is also interesting, as it specifically points to the role of calcium/phosphate homeostasis in the pathology. Rare variants in PTHLH are known to produce skeletal defects.^44^ A number of other genes involved in calcium/phosphate homeostasis, such as Klotho and FGF23 have been shown in animal model systems to develop kyphosis, likely through BMD loss and development of osteoporosis.^45^ As the PTHrP therapy is an approved treatment for osteoporosis, and is known to increase vertebral BMD, it might be possible to develop intervention strategies targeted to the kyphotic population with this genetic variation and potentially PTH deficiency in general.^46^

Other genes such as ETFA, critical to normal mitochondria maintenance and metabolism, could be contributing to the development of kyphosis via skeletal muscle declines.^47,48^ It is plausible that reduced capacity to repair cellular machinery in muscle and connective tissues, results in damage to these tissues over the lifetime of individuals, and consequently leads to increased spinal curvature.^49,50^ These effects are potentially exacerbated in individuals who carry mutations in cellular maintenance pathways and become more apparent in spine curvature as a function of age.^49,51,52^

Beyond genetic predisposition, our findings suggest that bone and muscle physiology are playing an important role in the progression of kyphosis. As such, interventions such as resistance training have the potential to improve both muscle mass and strength, as well as increase bone mineral density.^53–55^ Indeed physical therapy in hyper-kyphotic individuals was shown to improve kyphosis by ∼3-5°.^56–58^ It is also plausible that if increased spinal curvature is detected early, and can be attributed to osteoporosis, medications such as bisphosphonates could potentially offer additional benefits.^25,59^

Overall, our study identifies that declines in muscle and bone maintenance processes are greatly involved in age-related increases in spinal curvature. We show that increased body adiposity has distinct and opposing effects on lumbar and kyphotic angles, and increased spine curvature is associated with the development of GERD. Beyond age, maintenance of muscle mass is the key driver for kyphosis, while fat composition measures are the strongest drivers of lordosis. Analysis of genetic risk alleles suggests that molecular mechanisms involved are shared across multiple tissues and involve maintenance of cellular homeostasis such as DNA repair, RNA biogenesis, and mitochondrial functions.

### Limitations

There are several limitations of the current study. DXA scan used to quantify Cobb angles were conducted in the supine lateral position which could have a significant effect on spine angles as compared to an orthostatic measure.^60^ In the analysis of lumbar lordosis, we were not able to reliably identify L5 vertebrae due to pelvis obstruction, thus all Cobb angles were computed from L1-L4. The association between aging and spinal curvature was based on cross-sectional data, and thus does not represent individual aging trajectories, but rather overall population trends.

Participants of the UKBB imaging study are healthy volunteers recruited from the general community and thus tend to be healthier than the general population^61^; however, the cohort is well-suited to answer questions about various genetic, aging, and pathological factors that contribute to increased spinal curvature. Additionally, the significant colocalization findings of a few SNPs (rs9651832, rs3783820, rs9651832, rs71457747, and rs12908334) with RNA expression (eQTL/sQTL) of multiple genes may suggest a transgression of potential horizontal pleiotropy in the genetic analyses, especially for MR. Another limitation to note for the colocalization and MR analyses is that many skeletal and connective tissues are unavailable in GTEx. Thus, the possibility of associations between genetic expression in these missing tissues and spine angles cannot be dismissed.

## Supporting information

Supplemental Machine Learning Methods

## Acknowledgments

The authors would like to acknowledge Danny Park for his assistance with the bulk download of the EBI catalog. In addition, the authors would like to thank Elena Sorokin for downloading and processing GTEx v8 for colocalization and MR analyses. This research was supported by Calico Life Sciences LLC and conducted under UK Biobank Resource application number 18448.

## Methods

### Study population: The UK Biobank

The UK Biobank (UKBB) recruited 503,000 community-based adult volunteers aged 40-69 from 2006-2010 in 22 study centers across the United Kingdom: England, Wales, and Scotland.^62^ In the current analysis, we evaluate the UKBB imaging subgroup of 40,224 participants who returned between 2014-2020 to undergo spinal dual-energy X-ray absorptiometry (DXA) assessment (Lunar iDXA densitometer; GE Healthcare, Chicago, Illinois). At the baseline visit, extensive demographic and biometric information were collected including age, race, smoking status, height, and weight via questionnaires and physical measurements. The Townsend Deprivation Index (TDI), a composite measure for socioeconomic status calculated from unemployment, car and home ownership, and household crowding status, was evaluated from self-reported metrics.^63^ Additionally, blood was collected for serum biomarker and genomic analyses. Information on prior disease conditions were derived from baseline data and clinical data aggregated across healthcare providers: the Health Episode Statistics database from England, Patient Episode Database for Wales, and the Scottish Morbidity Record 01 for Scotland. The International Classification of Diseases, Tenth Revision (ICD10) was used.

This study was approved by the North West Multi-Centre Research Ethics Committee (MREC) and informed consent was collected from all participants. Further information on the study design of the UKBB has been previously published.^62^

### Machine learning

We constructed an image-processing pipeline that establishes a trace along the center of the vertebral column; then identifies an appropriate thoracic or lumbar section of the spine for analysis of kyphosis or lordosis, respectively; then measures the angle difference between lines tangential to the spinal traces at the top versus bottom of that section as a proxy for the Cobb angle. For the kyphotic angle, the thoracic spine approximately above and including vertebra T12 was analyzed; for the lordotic angle, the lumbar spine approximately between and including vertebrae T12 through L5 was analyzed. Full details of the method can be found in the Supplementary Method and GitHub repository (https://github.com/graham-calico/KyphoLordoDxa.git). Kyphosis and lordosis were measured using the python tool “spineCurve.py”. The following command-line options were invoked, for kyphosis: “--aug_flip --side_facing right”; and for lordosis: “--lumbar --aug_flip --side_facing right --aug_tilt 0.5”. Exact reproduction of the results in this manuscript should additionally invoke the “--legacy” flag. Further methods are described in the Supplementary Methods section.

### Statistical Methods

For all analyses, two-tailed P-values were calculated. Significance was defined as a false discovery rate-corrected p-value (pFDR) less than 0.05. All epidemiological analyses were performed with R version 4.3.1.

We assessed participant characteristics at the time of the imaging visit,, stratified for presence and absence of hyperkyphosis (>40°) and hyperlordosis (>30°). Continuous variables were summarized as medians and interquartile ranges (IQR), and categorical variables were presented as counts and percentages. The correlation between kyphotic and lordotic angles was displayed using locally smoothed regression with span=0.75 and the correlation coefficient (r) was calculated using Pearson product-moment correlation after confirming the linearity assumption could be reasonably met.

Unadjusted Wald unconditional maximum likelihood estimation was used to calculate the risk ratio between prevalent hyperkyphosis and hyperlordosis. To evaluate the association between age and kypholordotic angles, several analyses were conducted. First, the sex-stratified association between age and each kyphotic and lordotic angle was plotted using locally smoothed regression (span=0.75) with 95% confidence intervals (ggplot2). Second, participants were binned by age decile (e.g. 45-54, 55-64, etc.), and the proportion of individuals in each respective age category that had high kyphosis (40° to 50°+) and lordosis (30° to 40°+) angles by 5° intervals was graphed. Last, the mean age of participants in each 10° kyphosis by lordosis angle was presented as a heatmap.

To gain further insight into factors that might be contributing to increased spinal curvature, we used linear regression to evaluate the association of Cobb angles with 414 pre-existing medical conditions with prevalence >200 in our population, 59 clinical chemistry measures, and 15 body measure traits (adiposity, bone mineral density, muscle, lung traits) in UK Biobank. All associations were adjusted for age, age^2, sex, age*sex, smoking, BMI, and township deprivation index. FDR-adjusted significant associations between Cobb angle (either kyphosis or lordosis) and pre-existing conditions and clinical chemistry measures were presented. All 15 adiposity and skeletal-muscular body measure associations were presented, regardless of significance since they were related to our main study interest of phenotypic body composition associations with kypholordotic angle. All these skeletal-muscular associations were adjusted for variables listed above except BMI was not included in the adjustment of measures highly collinear with BMI calculation [BMI, height, total fat mass, and visceral adipose tissue].

We expected that the associations of various musculoskeletal traits on spine curvature could be non-linear. Thus, to assess ranked independent predictors of Cobb angles (out of body measure traits, age, sex, and smoking status), we ran variable selection using a 10-fold cross-validated boosted generalized boosted regression model with interaction depth = 2 and modeled selected features using splines in the generalized additive model (GAM).^64^ Variables with relative influence greater than 0 were selected for the model and relative variable influence percentages were presented. In secondary analyses, we reran all these linear regressions after stratifying by sex to examine potential effect modification by sex of these associations.

### Genetics

UKBB imputed genotypes were used in all genetic analyses.^15^ Further details on their methods have been previously published (https://biobank.ndph.ox.ac.uk/showcase/showcase/docs/impute_ukb_v1.pdf). We excluded SNPs with 1) minor allele frequency <1%, 2) low imputation quality with info value <0.9, 3) genotype missingness >10%, or 4) demonstrated deviation from Hardy-Weinberg equilibrium (HWE p<1e^−10^). Participants were excluded from the genomics analyses if they were 1) not of European ancestry (field ID 22006), 2) had heterozygosity or genotype call rate outliers (field ID 22027), 3) demonstrated sex chromosome aneuploidy (field ID 22019), 4) No mismatch between genetic sex and self reported sex (field ID 22001), and 5) Not genetically related (field ID 22011). This yielded a total of 9,911,384 SNPS and 33,413 participants in the genetic analyses. Bonferroni correction was used to evaluate significance in all the genetics analyses and a threshold of pFDR<0.1 was used. This study was conducted on a pre-existing dataset so was not preregistered.

### Genetic Wide Association Study

For the initial genetic wide association study (GWAS), a whole-genome regression model was conducted on kyphosis and lordosis separately using REGENIE, a computationally efficient, machine learning-based parallel analysis approach.^65^ In brief, REGENIE first uses cross-validated ridge regression by SNP block for dimension reduction of the genetic data. Then, a second cross-validated ridge regression for each trait is conducted to combine predictors from the first ridge regression into an overall prediction by trait, which is then decomposed by chromosome for a leave-one-chromosome-out (LOCO) scheme to decrease potential proximal contamination. These LOCO predictors are used as a covariate where each phenotype is tested by the set of imputed SNPs, also adjusting for genotype SNP chip (Illumina vs Affymetrix), sex, age, age^2^, age*sex, and recruitment center. We verified that the test statistics showed no inflation compared to the expectation using the genomic control lambda coefficient (1.13 for kyphosis and 1.09 for lordosis) and the intercept (0.999, SD 0.009 for kyphosis and 1.000, SD 0.084 for lordosis) of linkage disequilibrium score regression.

### Multi-trait Analysis of GWAS (MTAG)

The method for conducting MTAG from GWAS statistics has been previously described.^21^ MTAG enhances statistical power by leveraging the genetic correlation between traits to generate trait-specific estimates for each SNP. Based on linkage disequilibrium score regression (LDSC) estimates of genetic correlations between kyphosis and lordosis, a joint analysis of the two traits using the GWAS summary statistics was conducted using MTAG. Briefly, variants were restricted to those with minor allele frequency (MAF) > 0.01, and those that passed quality control (QC) in the individual trait GWAS. We verified that MTAG did not inflate the test statistics compared to the expectation using the genomic control lambda coefficient (1.14 for kyphosis and 1.11 for lordosis) and the intercept (0.998, SD 0.0085 for kyphosis and 0.9842, SD 0.0089 for lordosis) of LDSC.

### Genetic Correlation and Heritability

To examine if the genetic variation linked to spine angles shares genetics with other traits, we calculated genetic correlations with GWAS catalog phenotypes for kyphosis and lordosis, respectively. All available summary statistics from the EBI GWAS catalog (www.ebi.ac.uk/gwas/) were downloaded on October 19, 2022.^66^ Of the 25,475 summary statistics files, 20,021 could be successfully re-formatted by the munge step of LDSC.^67^ Of those, LDSC calculated a positive heritability for 14,843 summary statistics files. Of these traits, 10,170 and 10,138 for kyphosis and lordosis, respectively, yielded non-null SNP heritabilities and were used for downstream genetic correlation analysis.

In addition to traits available on the EBI GWAS catalog, we also estimated heritability of and genetic correlations between kypholordosis and musculoskeletal traits measured in the UKBB (adiposity, bone mineral content, muscle and lung traits). LDSC was calculated using the repository at https://github.com/bulik/ldsc/, version <u>aa33296</u>.

We estimated genetic correlation and heritability using default parameters and the –rg command and –h2 command, respectively (example: ldsc.py --rg kyphosis.sumstats.gz, lordosis.sumstats.gz --ref-ld-chr eur_w_ld_chr/ --w-ld-chr eur_w_ld_chr/ --out). Allele polarization was pre-harmonized to match the reference file w_hm3.snplist. Analyses were adjusted for sex, age, age^2^, age*sex, and recruitment center.

### Fine-Mapping

GCTA was used for approximate conditional analysis.^68,69^ We considered all variants that passed QC (above) and were within 500kb of the locus index variant, except the major histocompatibility complex (MHC) region due to the complex of LD structure of the region (GRCh38::6:28,510,120-33,480,577).^70^ For the LD calculation reference panel, we used genotypes from 5,000 randomly-selected, unrelated, Caucasian UKBB participants. For individual loci, variants with locus-wide evidence of association (p_joint_<1E-6) were considered conditionally independent. p<5E-8 was used as the genome-wide association threshold for defining a locus.

### Construction of genetic credible sets

For every distinct GWAS signal from, credible sets were calculated at a threshold of 95% probability of containing at least one variant with a true non-zero effect size.

First, the natural log approximate Bayes factor, Λ_j_, was computed for the *j*th variant within the fine-mapping region:

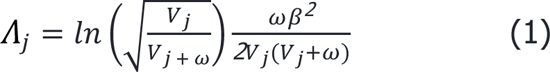

where β_j_ and V_j_ are the estimated effect size and corresponding variance, respectively.^71^ The parameter ω denotes the prior variance in allelic effects and is estimated as (0.15σ)^2^, where σ is the standard deviation of the phenotype. This standard deviation (σ) was estimated using the formula below:

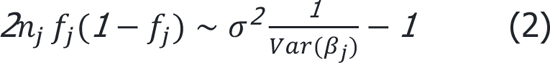

Here, Var(β_j_) is the variance of the beta coefficients, *f*_j_ is minor allele frequency, *n*_j_ is sample size, and σ^2^ is the coefficient of the regression which was used to estimate σ with σ = √σ^2^.

In genetic loci with multiple distinct association signals, we used two techniques 1) exact conditional analysis adjusting for all other index variants in the fine-mapping region and 2) marginal analysis not adjusting for other index variants within the locus. In genetic loci with only a single association signal, we used an unconditional meta-analysis.

Given *l* variants in the region, we then calculated the posterior probability, TT_j_, that the *j*th variant was driving the association using:

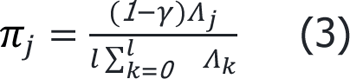

where γ denotes the prior probability for no association at this locus and *k* indexes the variants in the region (with *k*=0 indicating no association in the region). To account for the expected false discovery rate of 5%, we set γ=0.05 since a threshold of p_marginal_<5×10^−8^ was used to identify loci for fine-mapping.

For each signal, the 95% credible set was then constructed by (i) ranking all variants by their Bayes factor and (ii) including all ranked variants until their cumulative posterior probability exceeded 0.99.

### PheWAS

Fine-mapped lead SNPs were annotated to potential functional genes using the Ensembl Variant Effect Predictor, Open Targets, and dbSNP databases. We also considered previous significant associations in the GWASAtlas catalog and summarized our findings in **Table 1**.

### Colocalization

We performed colocalization tests to evaluate whether two traits (spine angle and gene expression) share the same underlying causal variants. For gene expression colocalizations, we used summary statistics from GTEx v8.^72^ For physiological and disease trait colocalizations, we used UKBB summary statistics of standardized quantitative and ICD10-categorized traits.^73^ We identified UKBB phenotypes where the minimum p-value within the +/-500kb region around the locus tag SNP was <5×10^-8^.

Colocalization analysis was conducted using the coloc R package with default priors and including all variants within 500kb of the index variant.^74^ Two genetic signals were considered to have strong colocalization evidence if PP3+PP4≥0.99 and PP4/PP3≥5 and suggestive colocalization evidence if PP3+PP4≥0.8 and PP4/PP3≥3.^74^

### Mendelian Randomization Analysis

To further evaluate causal associations between the genes identified driving spinal curvature, we conducted MR analyses.^75^ In MR analysis, we use cis-regulatory or splicing variants that affect RNA expression/function of implicated genes as instrumental variables and evaluate whether these expression and splicing changes drive changes in a phenotype of interest. Since genetic variants are theoretically randomly assigned when passed from parents to offspring, this method is expected to minimize the effect of confounding and avoid reverse causation.

The MR method relies on three important assumptions^76^:

(1) The genetic variant is associated with the exposure.
(2) The genetic variant is not associated with any confounders.
(3) The genetic variant is not associated with the outcome through any pathway other than via the exposure (no horizontal pleiotropy).

Since multiple tissues can affect spine curvature, we carried out two sample MR analyses across all 54 tissue types measured in GTEx v8.^72^ To identify independent SNP instruments for each exposure, we pruned GWAS-significant SNPs (p-value < 5 × 10^-8^) for each risk factor using a threshold of r^2^ < 0.01 and LD window of 250 kb. The construction of reference panels for LD calculations was described in the fine-mapping section. When multiple independent eQTLs were observed for the same gene in a single tissue within GTEx, we evaluated the causal effect of a gene within each genome-wide significant locus on kyphosis and lordosis using inverse variance weighting (IVW). For genes with only a single independent eQTL association in a tissue, we used the Wald ratio test. A notable limitation of eQTL-based MR analysis is that bone and connective tissues are not available in the current version of GTEx (v8), and we cannot rule out the possibility that causal effects on spine angles are not due to gene expression in missing tissues.

For sensitivity analyses, the MR analysis was also reconducted using 1) MR-Egger regression and 2) weighted median-based and mode-based tests of the ordered Wald ratio estimates.^77,78^ Since weighted median and mode estimates assumes that the estimates from pleiotropic variants are outliers, these methods tend to be less sensitive to the effect of pleiotropic variants.^79^ On the other hand, MR-Egger involves a weighted linear regression to evaluate the marginal effect of each SNP to the outcome on the marginal effect of each SNP to the exposure. Thus, this method evaluates the overall directional pleiotropic contribution of weak instrumental SNPs on the risk estimate. As such, we used these methods as sensitivity analyses to confirm that there was a consistent directional effect between the variants and traits as from the IVW and Wald ratio results.^79^

### Ethics aspects

All UKBB data used in this study were anonymized and were accessed through project number 18448. The National Research Ethics Service Committee of North West-Haydock (REC reference: 11/NW/0382) approved the UKBB project. Signed informed consent was obtained electronically from all participants https://biobank.ctsu.ox.ac.uk/crystal/crystal/docs/Consent.pdf.

## Data availability

Data from the UK Biobank is available on request (https://www.ukbiobank.ac.uk/). Summary statistics are available from the GWAS catalog under accession number (TBD).

## Code availability

Our open-source code can be accessed at https://github.com/graham-Scalico/KyphoLordoDxa.git.

