## Supplemental Machine Learning Methods for "Aging of the Spine: Characterizing genetic and physiological determinants of spinal curvature"

**Machine Learning - Computational pipeline for kyphosis/lordosis measurement 1**
Step 1: Determination of whether the DEXA image is left-facing or right-facing. 1
Step 2: Segmentation of the body of the spine. 2
Step 3: Identification of thoracic and lumbar regions. 2
Step 3a: Identification of anterior intervertebral junctions. 2
Step 3b: Definition of thoracic and lumbar regions using intervertebral junctions. 3
Step 4: Geometric estimation of the curvature of the spine in each designated region. 4
**References 8**

### Machine Learning - Computational pipeline for kyphosis/lordosis measurement

The machine learning pipeline described here automates the angular scoring of curvature either in the upper/thoracic spine (kyphosis) or lower/lumbar spine (lordosis), using a lateral dual-energy X-ray absorptiometry (DXA) scan image of a human torso. These angular values that are derived estimate Cobb angles in the two spinal regions which are the most commonly used measurement for quantifying spinal curvature.[1]

The analysis occurs in four steps:

1. Determination of whether the DEXA image is left-facing or right-facing;
2. Segmentation of the body of the spine;
3. Identification of anterior intervertebral junctions;
4. Geometric estimation of spine curvature in the designated region.

Details on each of those steps are given below, along with the final performance of the system against hold-out test data generated by human annotators.

#### Step 1: Determination of whether the DEXA image is left-facing or right-facing.

The sign of the observed angle for either kyphosis or lordosis depends on the orientation of the image and a conventional (but geometrically arbitrary) decision about which direction of rotation to define as positive. For this application, zero angle is defined as a perfectly straight spine, and positive angles are defined based on the typical curvature of the spine, when facing forward: concave for kyphosis and convex for lordosis.

By default, this application uses an image classification model to determine whether an image is left-facing or right-facing. That model is a re-trained version of the ResNet V2 50 model, originally trained for imagenet (ILSVRC-2012-CLS) classification.[2,3] This model achieved 100% accuracy for correct sorting of 91 images from UK BioBank, each presented as found in the UKBB (right-facing) or horizontally flipped (left-facing) - i.e. 182 test images.

Given the consistently right-facing orientation of side-angle DEXA scans in the UKBB, a command flag was implemented to force analysis assuming all images to face in a given direction, called as either --side_facing right or --side_facing left.

#### Step 2: Segmentation of the body of the spine.

In order to trace the curvature of the spine, a semantic segmentation model was trained to label the body of the spine. The body of each vertebra was labeled, along with the spaces in between. This model was trained using the tool provided by the "image-segmentation-keras" code base (https://github.com/divamgupta/image-segmentation-keras commit f04852d from September 6, 2019), specifying the "vgg_unet" model architecture[4], with input widths and heights of 192 and 384, respectively. This model labeled a manually-annotated test set of 59 images with a mean IoU accuracy of 0.889 (SD 0.021).

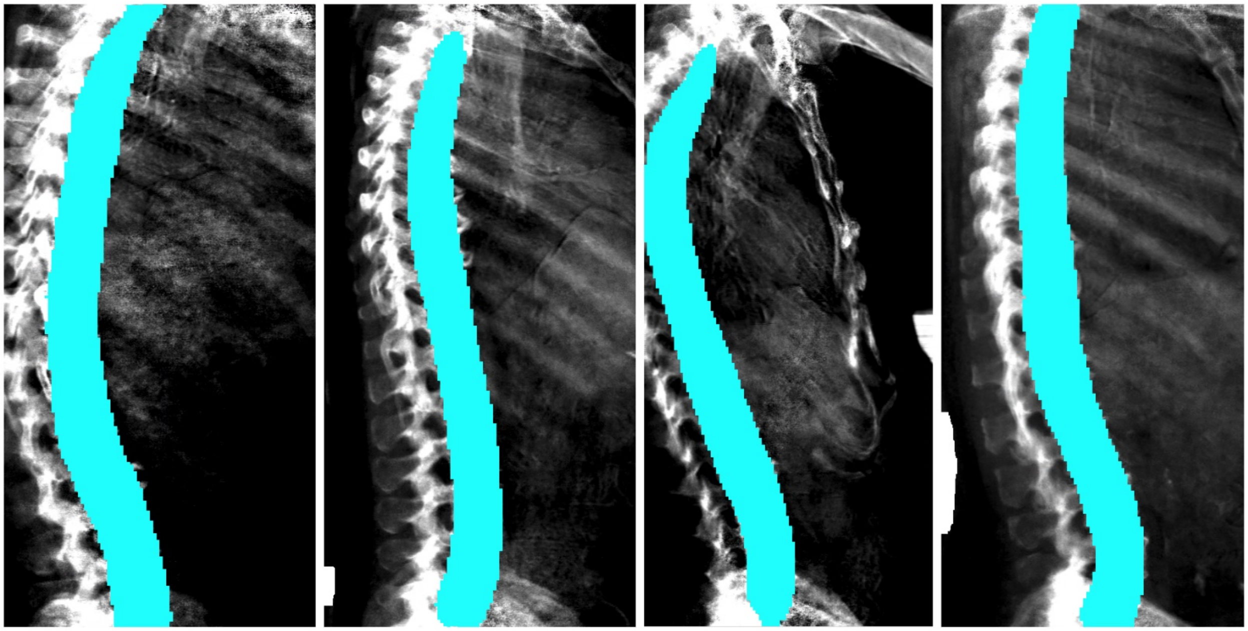

#### Step 3: Identification of thoracic and lumbar regions.

##### Step 3a: Identification of anterior intervertebral junctions.

In order to register the position along the spine, to determine the appropriate portion of the spine along which to measure curvature, the anterior edge of each intervertebral space is identified using an object detection model:

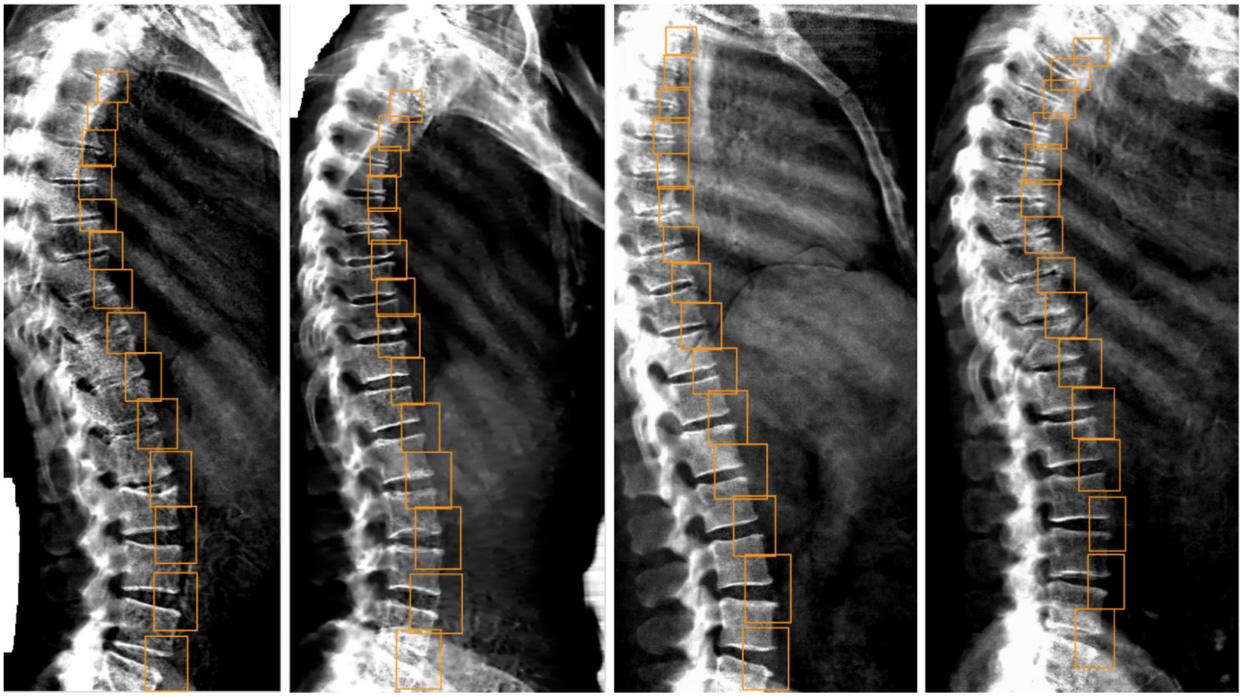

This model was trained for the ML measurement of DISH (https://github.com/calico/DISH).[5] Its performance and details of its training are described in that repo. As for DISH, the 14 top-scoring predicted boxes are used for downstream analysis.

##### Step 3b: Definition of thoracic and lumbar regions using intervertebral junctions.

For both left-facing and right-facing images, it was always assumed that the image was in an upright orientation (i.e. the person's shoulders were at the top and pelvis was at the bottom of the image). It was also assumed that each image was of the spine from approximately the shoulders to the hips. Therefore, for both kyphosis and lordosis, curvature was measured from the cut-off point derived below all the way to the appropriate edge of the image (top edge for kyphosis, bottom edge for lordosis).

The center points for each of the 14 intervertebral disc boxes generated were calculated, then sorted along the y-axis, from top to bottom. The y-axis cut-off point was defined using the y-axis values from that sorted list of coordinates. For kyphosis, the cut-off was set to halfway between the 11th and 12th (zero-indexed) y coordinates from the list. For lordosis, the cut-off was set to 25% of the distance between the 10th and 11th y coordinates from the list.

For kyphosis, the region of interest was defined as being above the cut-off, i.e. all the way to the top of the image. For lordosis, the region of interest was defined as being below the cut-off, with a further refinement that is described under Step 4 - i.e. almost all the way to the bottom of the image.

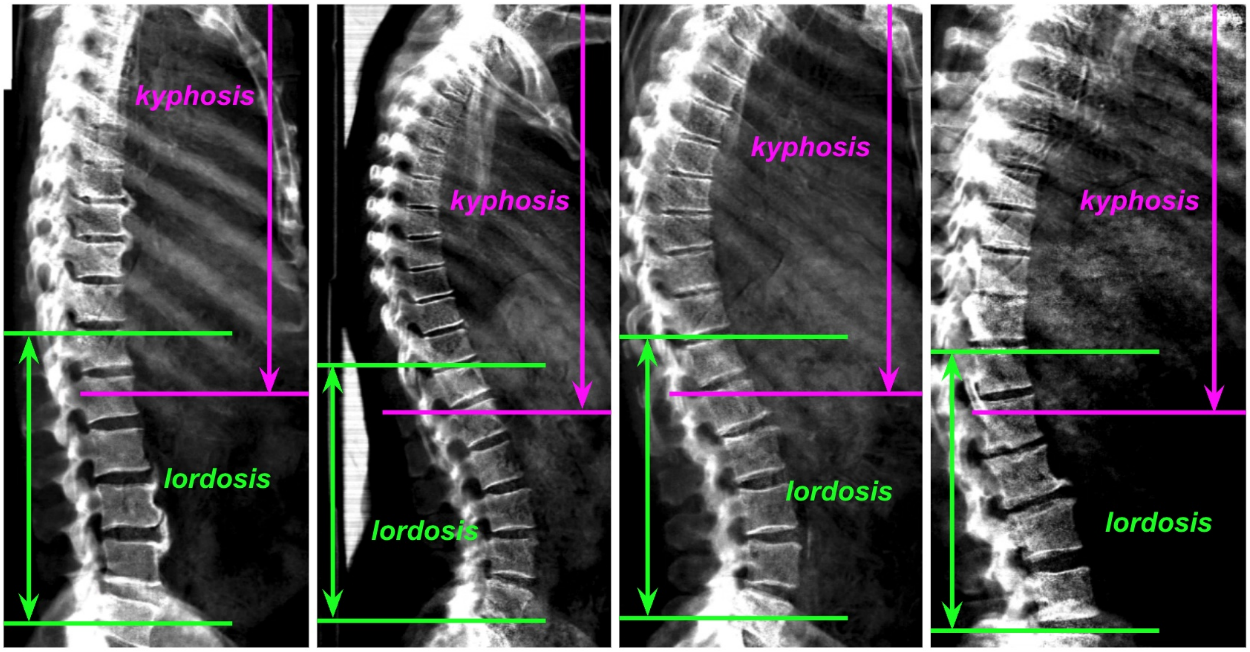


#### Step 4: Geometric estimation of the curvature of the spine in each designated region.

In order to define a consistent reference axis across images (the spine axis), an orthogonal distance regression (ODR) was taken through the spine mask (shown below in yellow), counting each masked pixel as a data point. The mask pixel points were then sorted along that axis. A spinal trace was generated by dividing that sorted list of points into 90 bins. For each bin, the average position of all points in the bin was taken, and that value was added to the spinal trace (shown below in red).

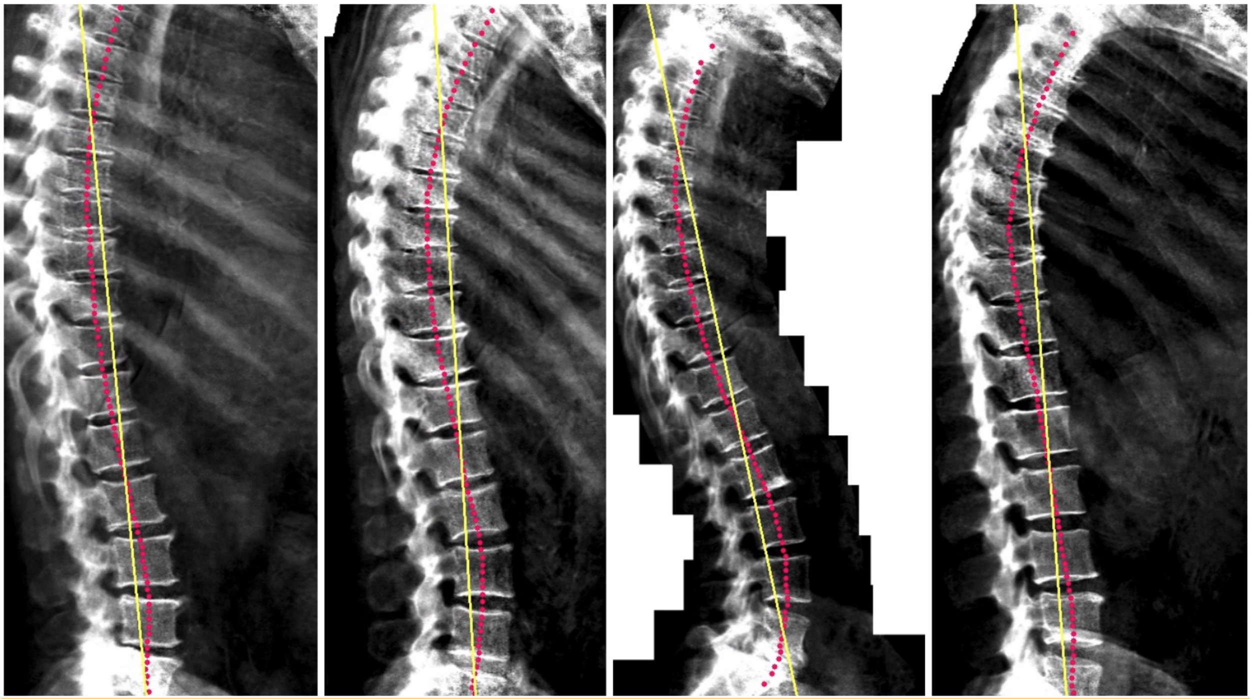

At this point, for lordosis, the region of interest was refined at the bottom by excluding the bottom-most 6% of data along the spine axis.

Shown below: for each analysis (kyphosis or lordosis), a new spine axis was then calculated using ODR on the spinal trace points that fell within the region of interest. These new axes are shown below, in pink for kyphosis and in green for lordosis. The new axis was then used to re-compute the spinal trace using the same procedure as above (and shown below as dots matching the colors of their corresponding axis). Note that the original implementation of this software did not re-compute the trace. Though it had negligible impact on the output, that original behavior can be invoked using the --legacy flag.

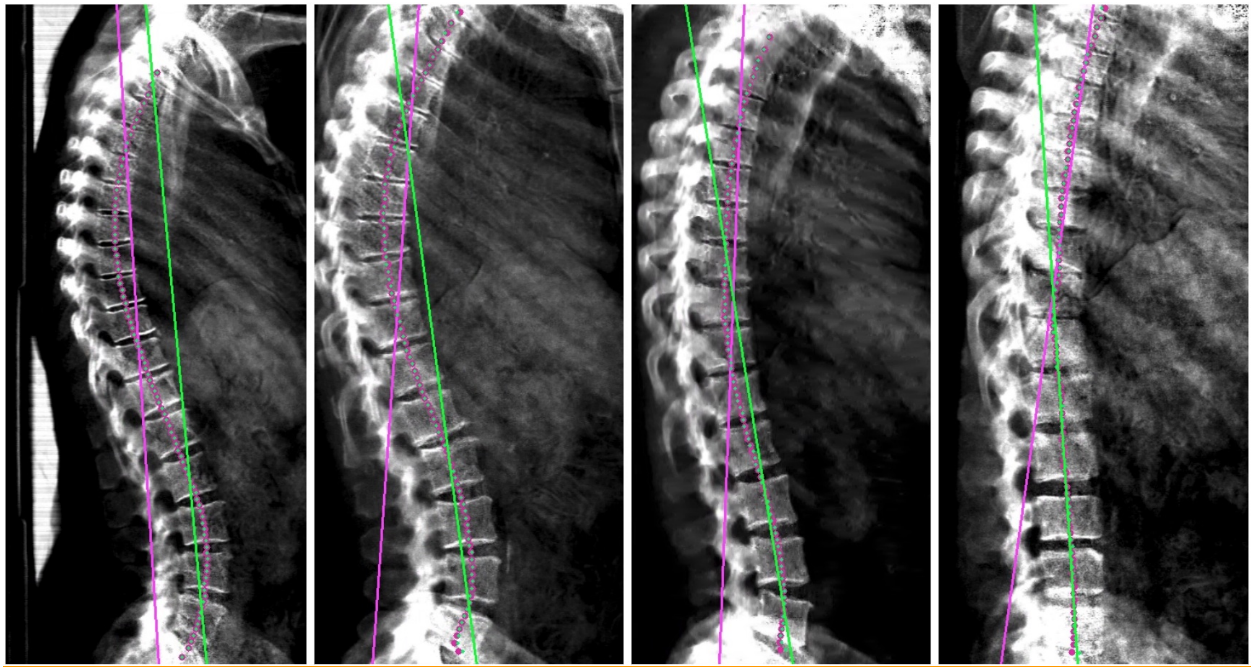

To measure spinal curvature, spinal trace points whose positions along the spine axis fell within the region of interest, and sorted along that axis, were divided into three equally-sized sets. ODR lines were taken for the spinal trace points from the sets highest and lowest along the axis. Below, the relevant points are in red, and their derivative vectors are in yellow.

Shown for kyphosis:

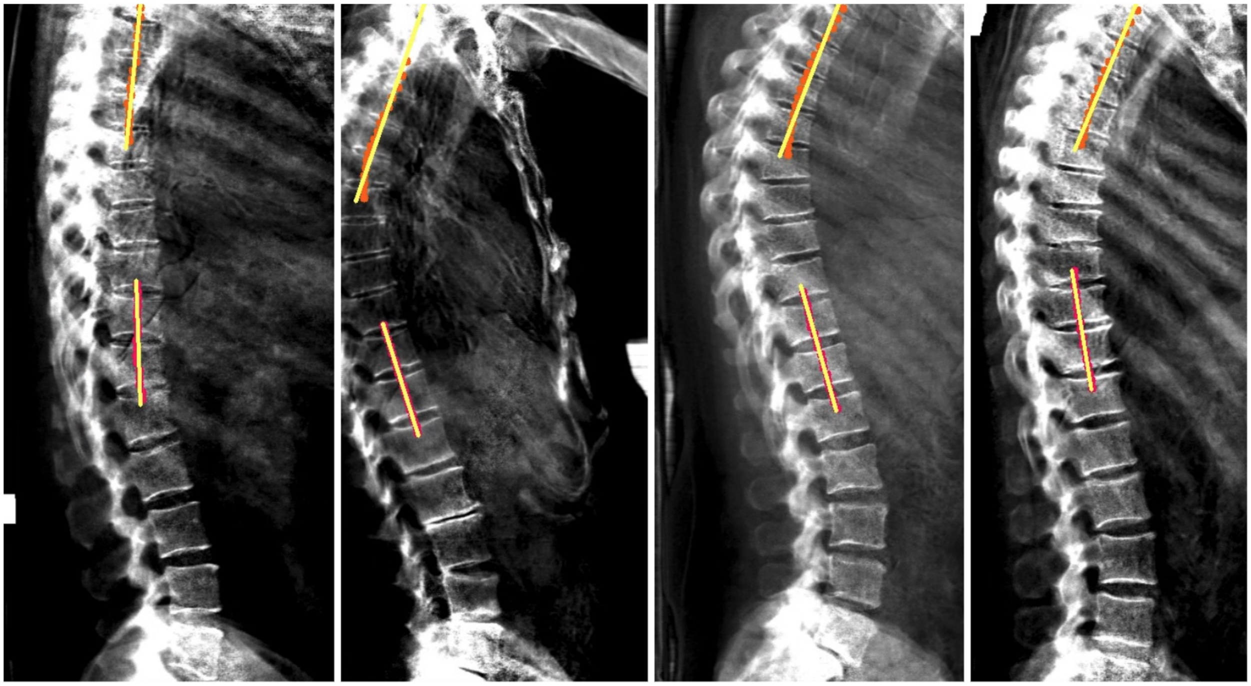

Shown for lordosis:

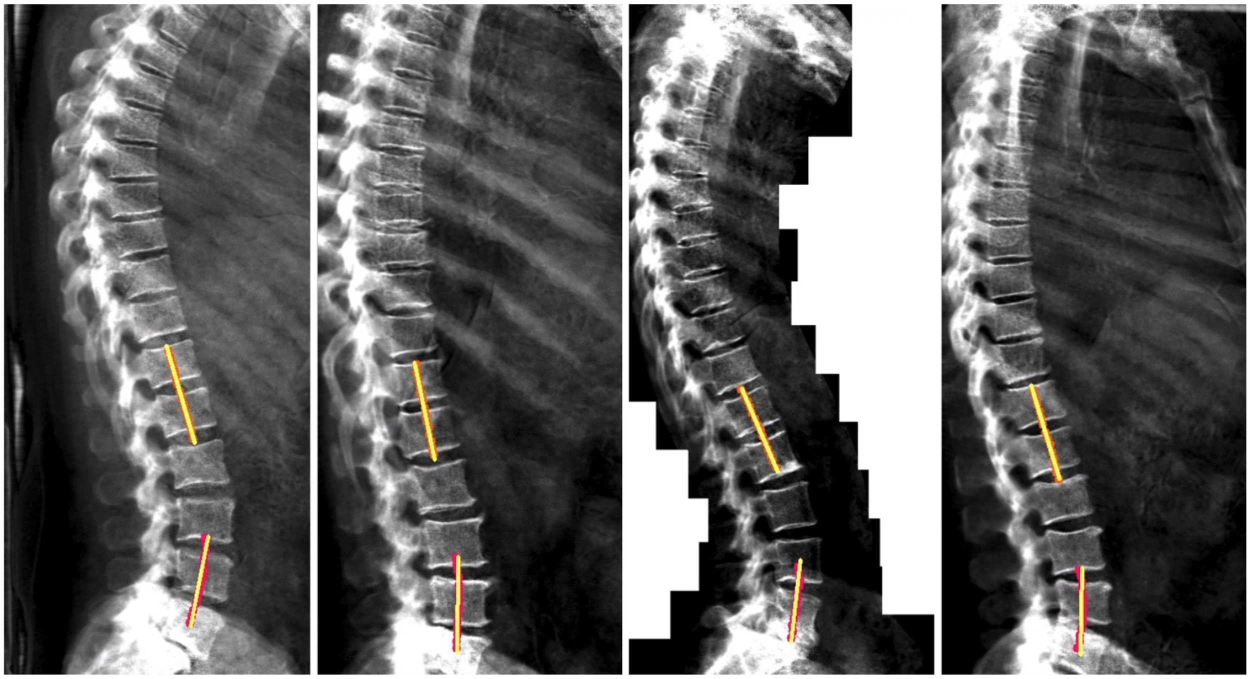

The angle difference between those two lines was reported as the Cobb angle estimate with sign adjustment, as described above.
