## Supplemental Figures for "Aging of the Spine: Characterizing genetic and physiological determinants of spinal curvature"

### Supplementary Figures

Supplementary Fig 1. Validation of annotator and ML model concordance 1
Supplementary Fig 2. Correlation between kyphosis and lordosis angle 2
Supplementary Fig 3. Sex-stratified comorbidity and spine curvature association 3
Supplementary Fig 4. Nonlinear comorbidity and spine curvature association 4
Supplementary Fig 5. Sex-stratified biochemistry and spine curvature association 5
Supplementary Fig 6. Nonlinear musculoskeletal and spine curvature association 6
Supplementary Fig 7. Adiposity and spine curvature association 7
Supplementary Fig 8. Sex-stratified musculoskeletal and spine curvature association 8
Supplementary Fig 9. GWAS (non-MTAG) with fine-mapping for spine curvature 9
Supplementary Fig 10. PheWAS of significant loci 10

**Supplementary Figure 1. Annotator and ML Model Concordance on Validation Set for Lordosis and Kyphosis Cobb Angles.** H1-H4 are human annotators. H.Mean = Annotator Mean Angle, ML = Machine Learning Cobb Angle. Correlation coefficients are shown in the upper triangles.

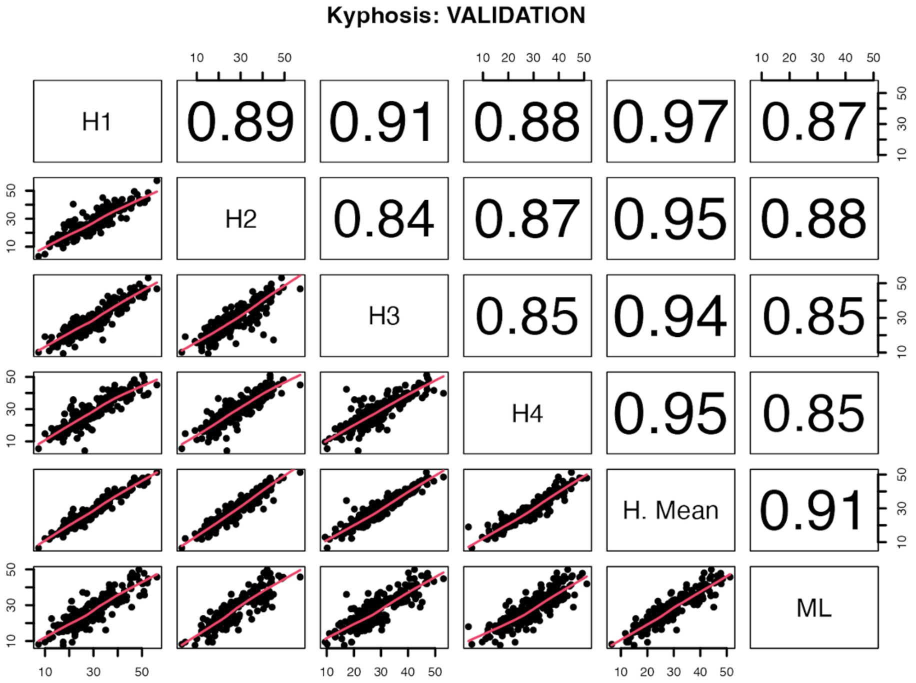


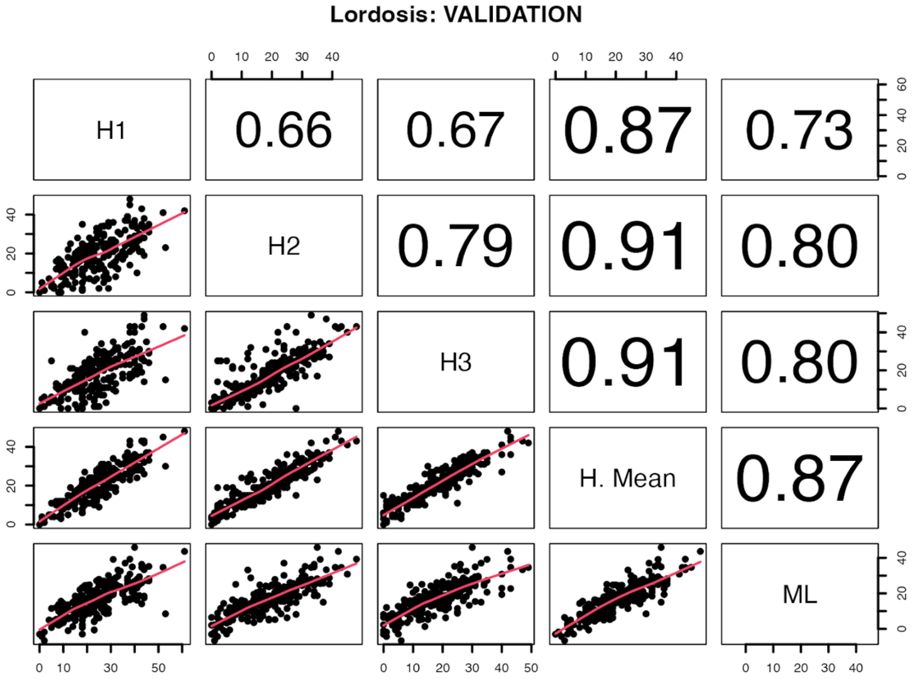

**Supplementary Figure 2. Correlation between kyphosis and lordosis angle.** The blue line indicates the non-linear loess positive association between the two traits. Gray shading is the 95% confidence interval of the nonlinear association.

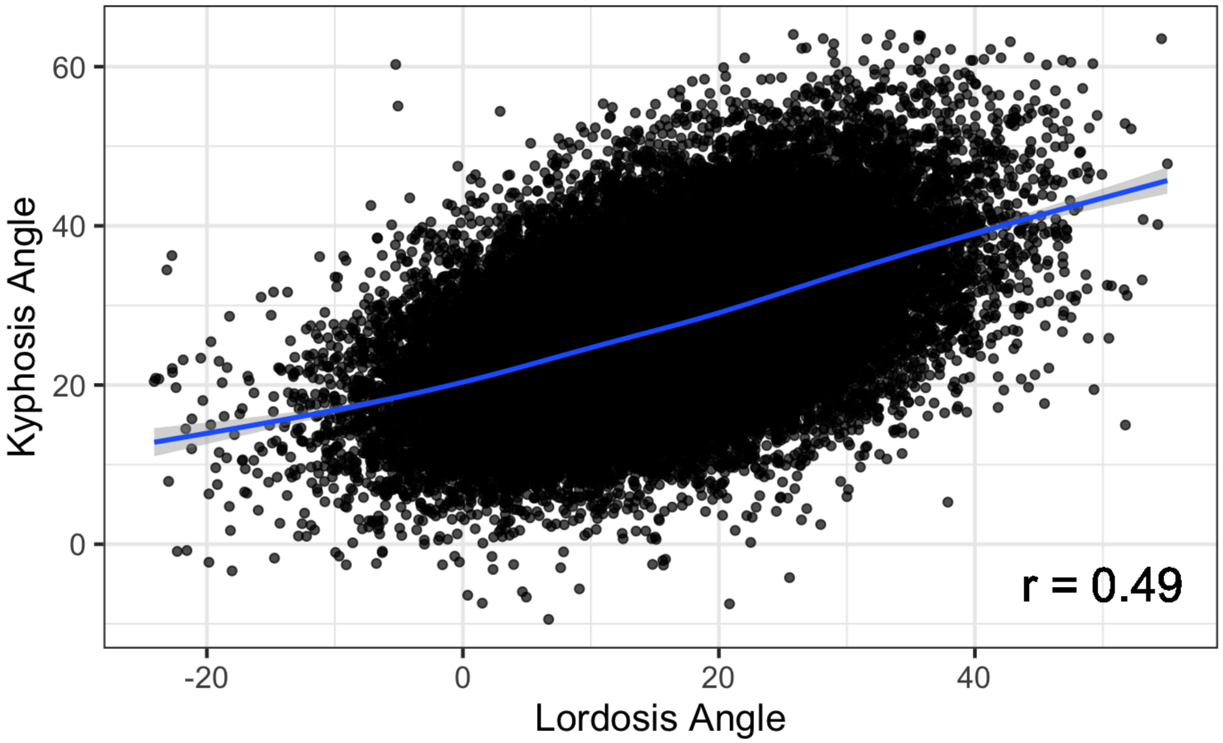

**Supplementary Figure 3. Sex-stratified associations between prior diseases and kyphotic/lordotic angle.** All analyses were adjusted by age at visit, BMI, smoking status, and Townsend deprivation index.

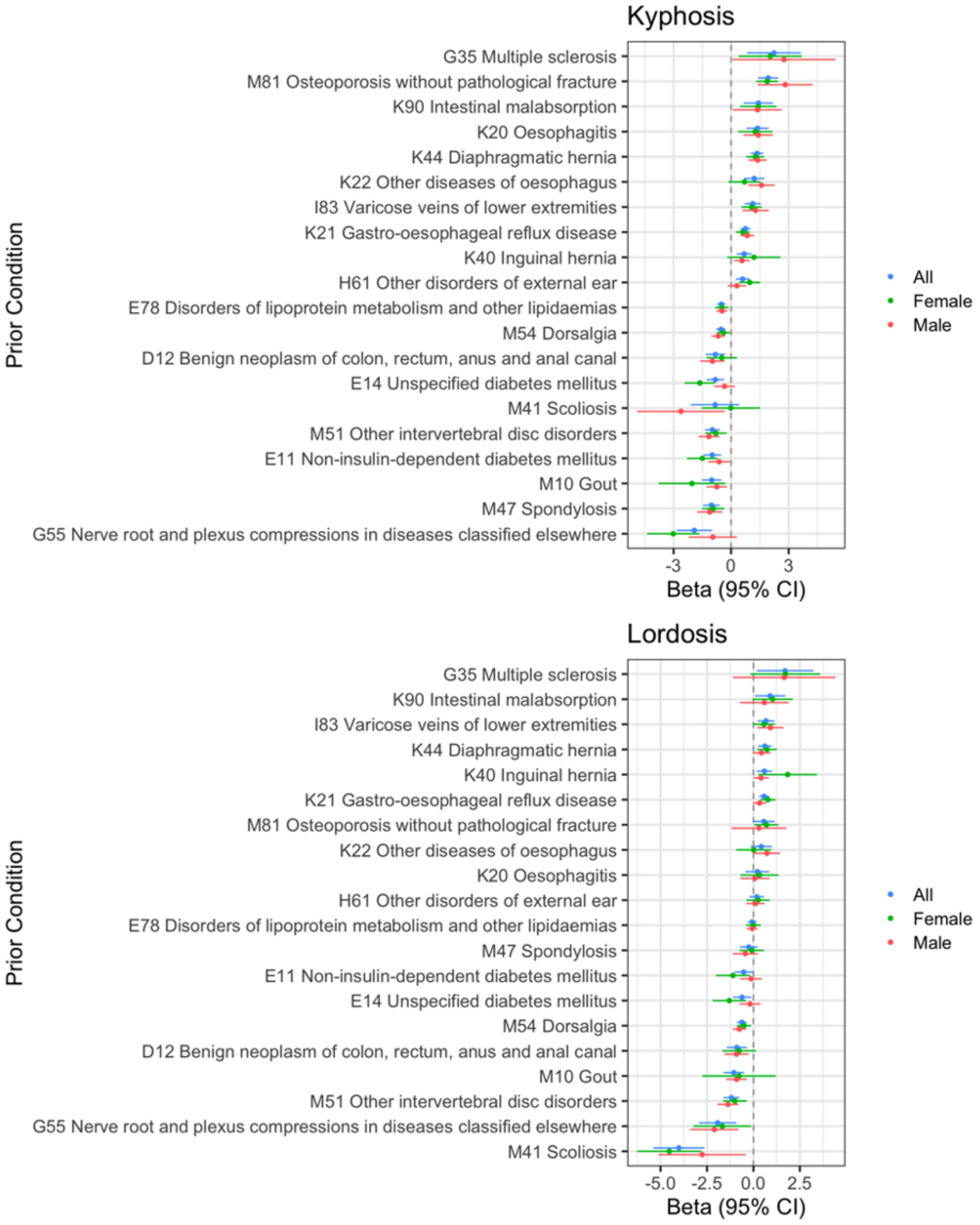

**Supplementary Figure 4. Nonlinear associations between kypholordotic angles and adjusted probabilities of pre-existing comorbidities A) Positive associations B) Inverse associations.** Red star annotations indicate significant association in adjusted linear regression models. Analyses adjusted by age at visit, BMI, smoking status, and Townsend deprivation index.

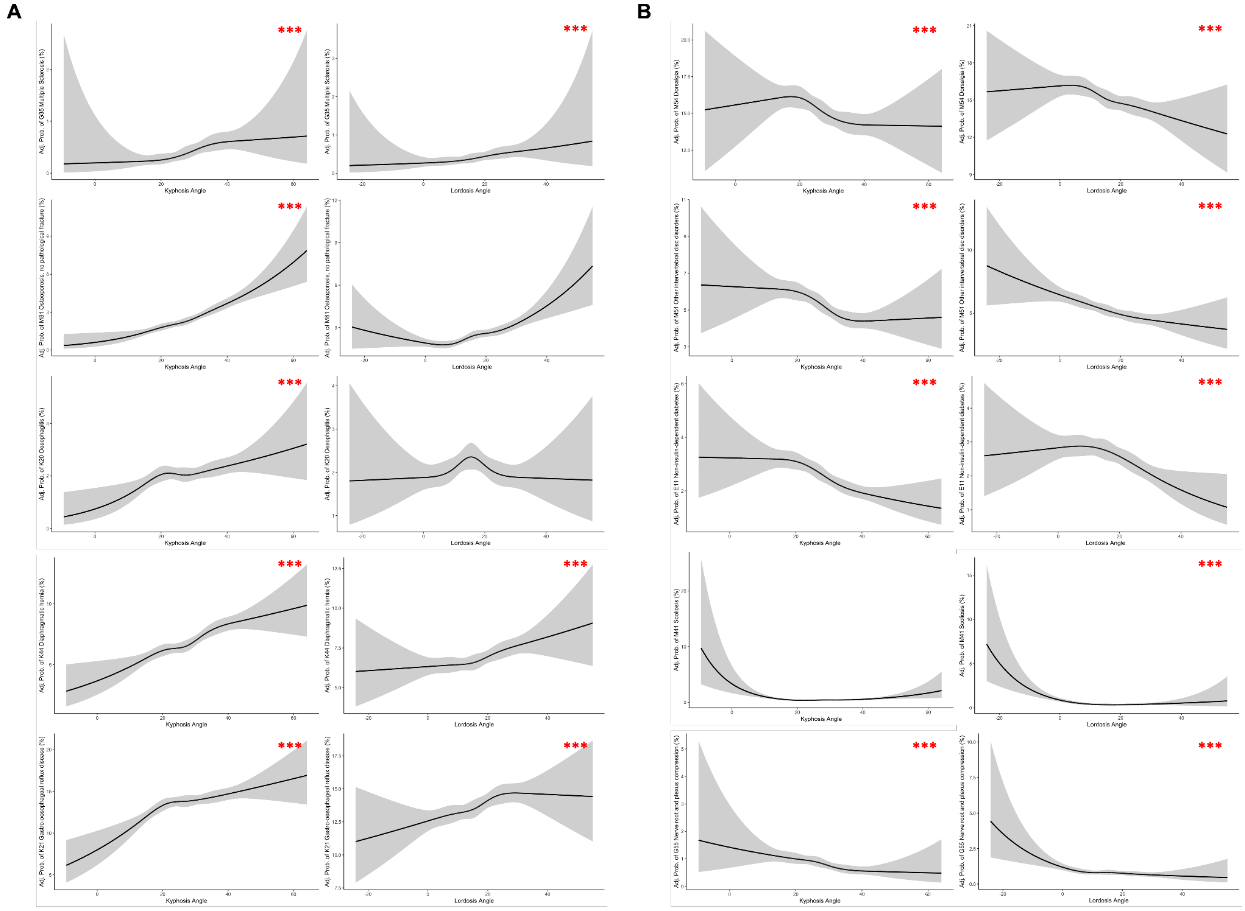

**Supplementary Figure 5. Sex-stratified associations between serum biochemistry and kyphotic/lordotic angle.** All analyses were adjusted by age at visit, BMI, smoking status, and Townsend deprivation index.

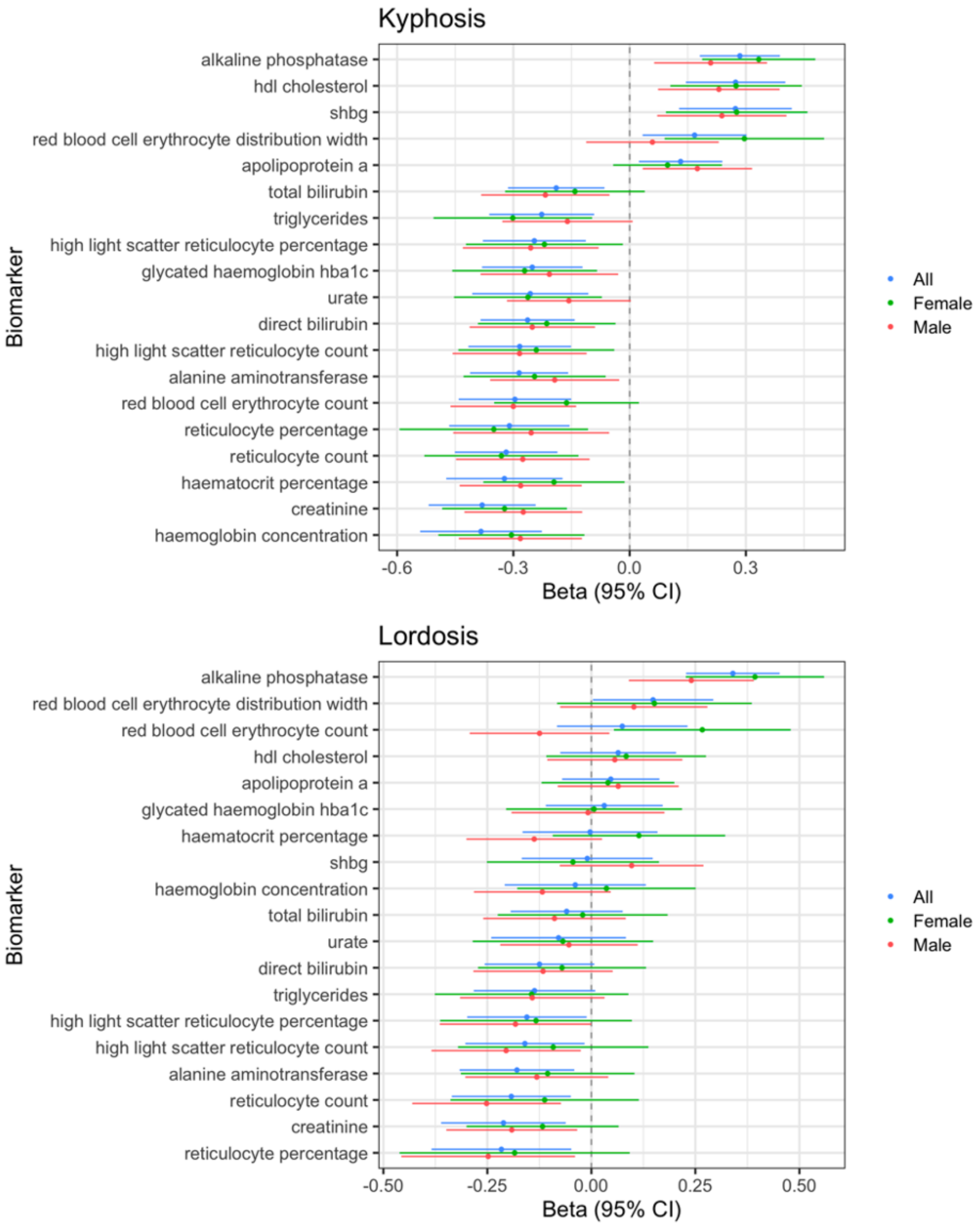

#### Supplementary Figure 6. Non-linear sex-stratified correlations between musculoskeletal traits with kyphosis and lordosis angles.

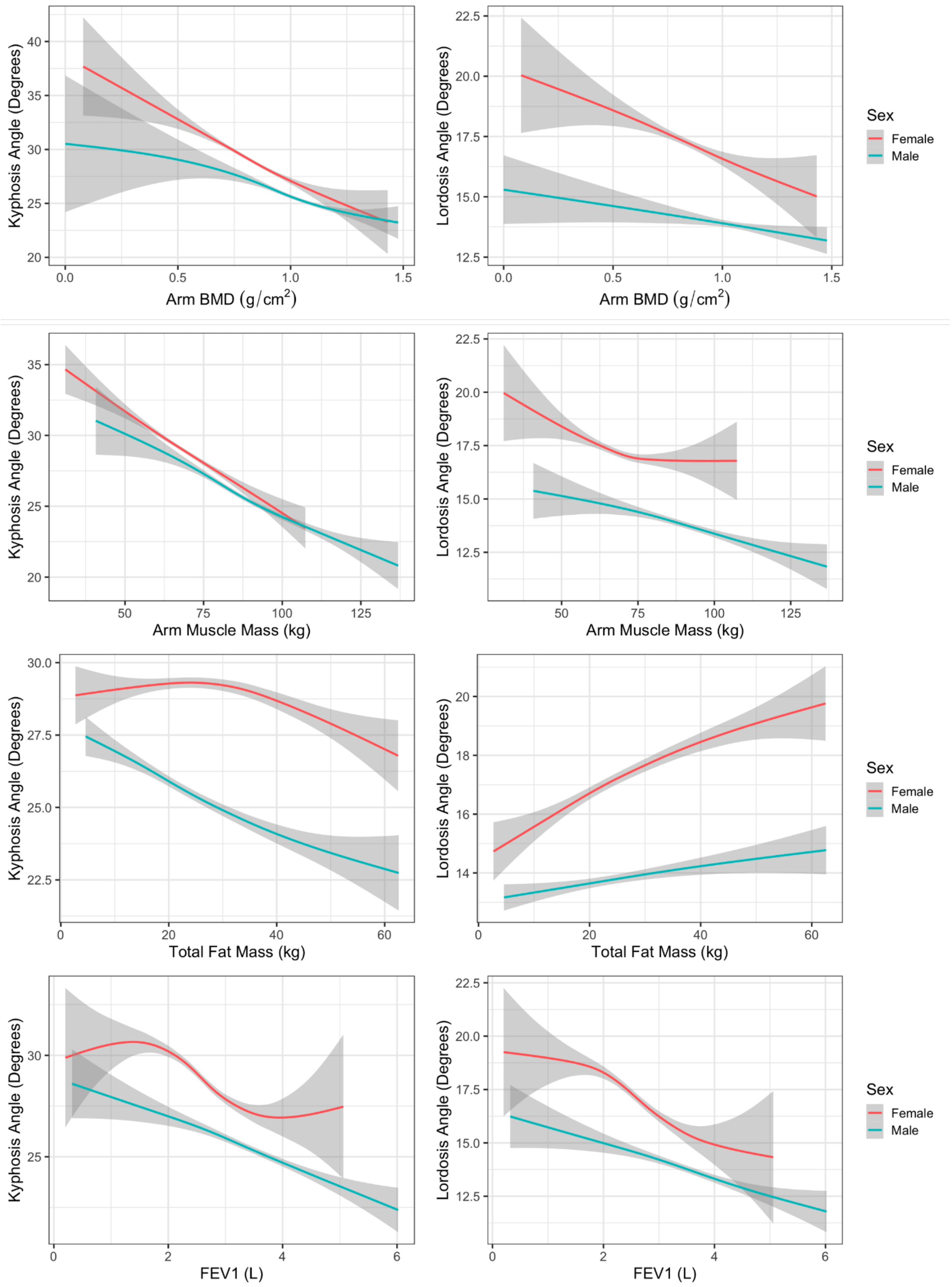

**Supplementary Figure 7: Association between adiposity and kypholordotic angles.** Adjusted models of kyphosis and lordosis accounting for age, respective angle, scale Townsend deprivation index, and scaled total fat.

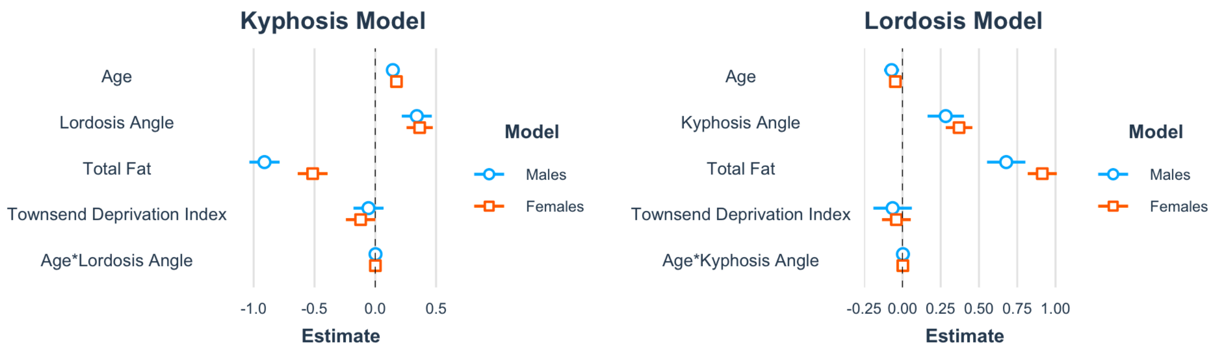

**Supplementary Figure 8. Sex-stratified associations between musculoskeletal traits and kyphotic/lordotic angle.** Analyses were adjusted by age at visit, sex, BMI, smoking status, and Townsend deprivation index, except BMI, height, total fat mass, and visceral adipose tissue were not adjusted for BMI.

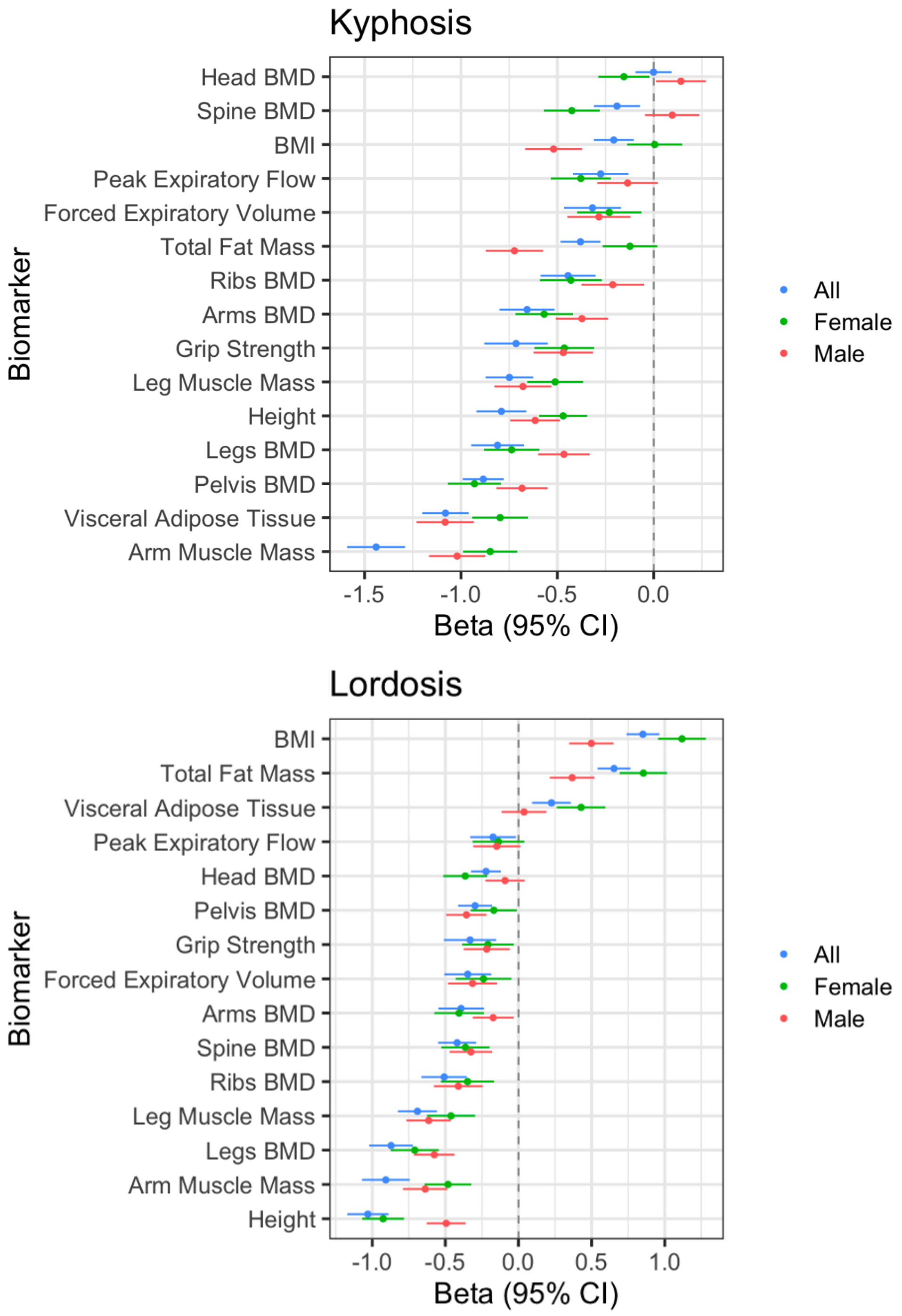

**Supplementary Figure 9. Non-MTAG GWAS for kyphosis and lordosis with fine-mapped lead SNPs annotated.**

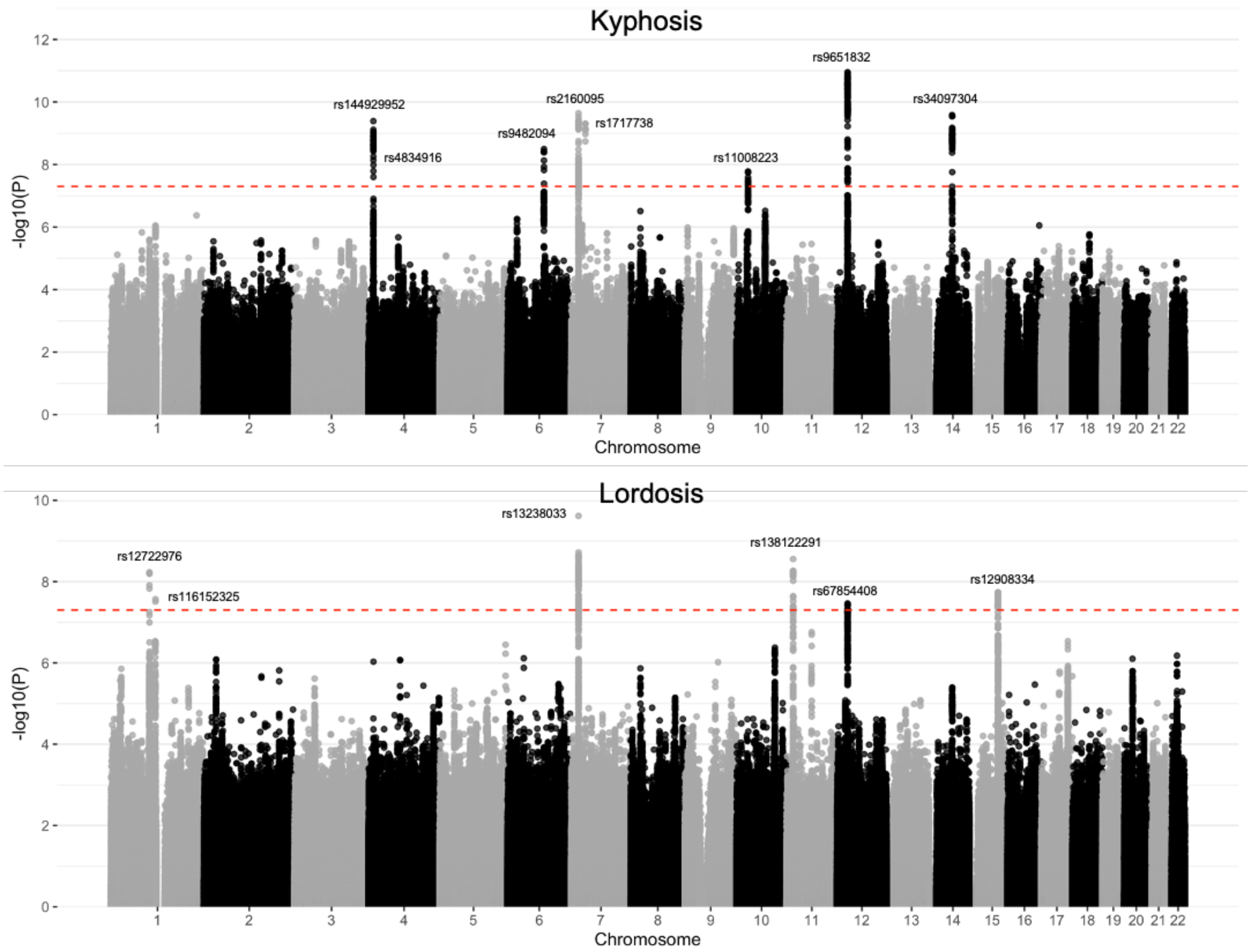

**Supplementary Figure 10. PheWAS plots of significant kypholordosis loci from the GWASAtlas.** Loci identified by GWAS were associated predominantly with skeletal, respiratory and metabolic traits. Only SNPs with significant trait associations (p<5E-8) are displayed.

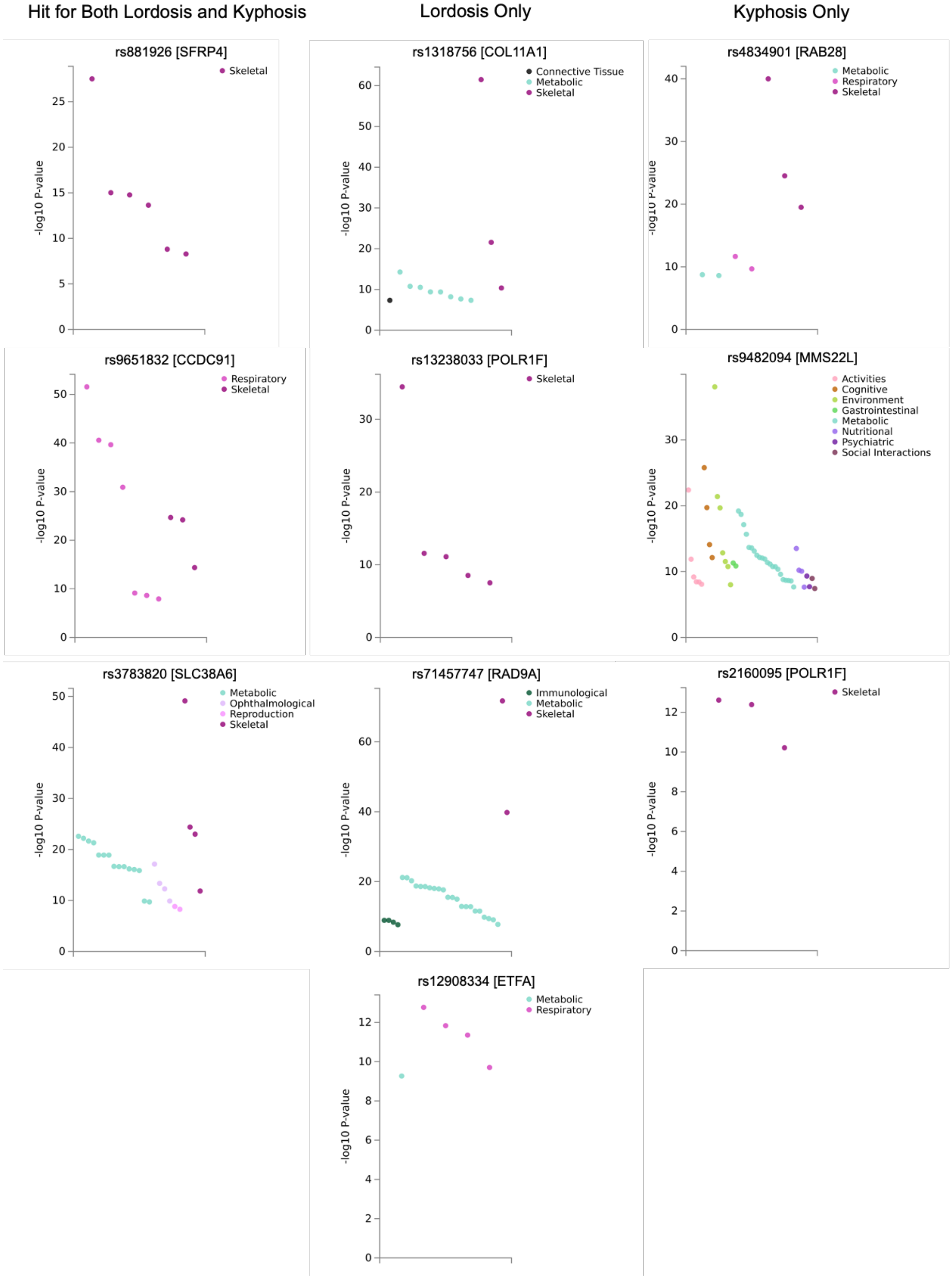
